# Kinetic modeling of transaminase values distinguishes active liver inflammation from resolution independent of absolute enzyme levels

**DOI:** 10.1101/2022.09.14.22279084

**Authors:** Marc S Sherman, Wolfram Goessling

## Abstract

The biomarkers alanine (ALT) and aspartate (AST) aminotransferase serve an indispensable role in the diagnosis and management of liver diseases where patients often have no symptoms until organ function is threatened. Despite 70 years of widespread clinical use, the exact kinetic behavior of AST and ALT during liver injury in humans has never been quantified. Here, we used mathematical modeling to examine the individual trajectories of > 6.5 million AST and ALT levels over time in 91,086 patients with acute liver injury. Candidate mechanistic differential equation models fitted to these trajectories revealed that 40.1% of ALT and 37.5% of AST curves were well-fit by a single-exponential model, indicating a single rate-limiting step governs transaminase decline. The mechanism of this rate-limiting step was found to be transaminase clearance from the blood, rather than ongoing liver injury, highlighting that AST and ALT are lagging biomarkers of liver injury. We use this observation to infer the plasma clearance rate of AST (t_1/2_=15.8h) and ALT (t_1/2_=34.6h) in humans, and derive a calculator of real-time liver injury, the Hepatocyte Injury indeX HIX (hix.massgeneral.org) that accounts for lag due to clearance of serum transaminase. We demonstrate that this index distinguishes active from resolved liver injury on histopathology assessment, even when the absolute AST or ALT value remains markedly elevated. These observations sharply refine how transaminase patterns are interpreted in the practical care of patients with acute liver injury.

**One Sentence Summary:** Kinetic modeling of >6 million individual AST and ALT values from 91,000 patients enables biomarker half-life estimation and a real-time liver injury assessment.

## Introduction

Clinicians have diagnosed and monitored liver disease using the biomarkers alanine aminotransaminase (ALT) and aspartate aminotransaminase (AST) since 1955 *(1)*, and both tests remain among the most commonly ordered laboratory panels in clinical medicine *(2)*. The underlying assumption, taught to trainees at every stage of their medical education, is that the level of abnormal transaminases corresponds proportionally to the level of active liver injury, where higher levels indicate worse injury *(3–5)*. Transaminase levels and their trajectory over time help clinicians decide when to obtain a liver biopsy *(5, 6)*, initiate and monitor response to therapy *(5, 7–9)*, refer for liver transplantation *(10)*, or discharge a patient from the hospital.

Despite the essential role of interpreting liver enzymes in the management of patients with liver disease, there has been no quantitative characterization of the kinetic behavior of AST and ALT in a large clinical cohort. This leaves open the possibility that clinical decision making based on transaminase pattern could be considerably refined in the same way that that creatinine kinetics anticipate acute kidney injury *(11)* and glycation kinetics improve hemoglobin A1c estimates *(12)*. In particular, the plasma clearance rate is an essential kinetic property of all biomarkers that determines how quickly biomarkers like transaminases respond to underlying organ injury *(13)*. A biomarker that is slowly cleared (long half-life) responds slowly to acute changes but remains positive after injury resolves and is therefore best suited for marking a missed event. A biomarker with a fast clearance rate (short half-life) is highly responsive to acute changes, but rapidly disappears, and is therefore a more faithful reflection of the current state of injury. Clinically, transaminases are treated as a biomarker with fast clearance, though it is not known where AST or ALT falls on this spectrum in humans.

Here we study the kinetics of AST and ALT during acute liver injury with a focus on the plasma clearance rate of transaminases in humans. We used mechanistic mathematical models to quantitatively characterize the transaminase recovery patterns in 91,086 patients comprising more than 6 million AST and ALT measurements across a multi-hospital health system (Mass General Brigham) from 1995 to 2021, agnostic to underlying disease. We illuminate how different model-fits in single-patient encounters leads to estimation of the fundamental plasma clearance rate for AST and ALT in humans and validate hypotheses by comparing model prediction with gold-standard liver pathology. Finally, we show how the plasma clearance rate of transaminases enables construction of a real time hepatocyte injury index (hix.massgeneral.org) which is immediately deployable to clinical decision making.

## Results

### Mathematical modeling indicates AST and ALT recovery is often defined by first-order kinetics

A query for patients with acute liver injury defined by at least one ALT>150 IU/L and >6 total measurements of ALT yielded a database of 91,086 patients comprising 3.2 million ALT results and 3.3 million AST results. Individual patient trajectories were extracted using a custom Python pipeline (Fig. 1A). For each trajectory, peaks and troughs (Fig. 1A-D) were annotated using SciPy’s signal processing package *(14)*. Individual transaminase peaks correspond to clinical episodes of liver injury. To understand whether an episode might be amenable to modeling, we determined how many laboratory measurements were made before and after each peak. This revealed that laboratory testing was heavily biased post-peak, compared to pre-peak, at a ratio of 2:1 or greater (Fig. 1E). We therefore focused our initial efforts on understanding the kinetics of post-peak recovery trajectories.

**Figure 1.**
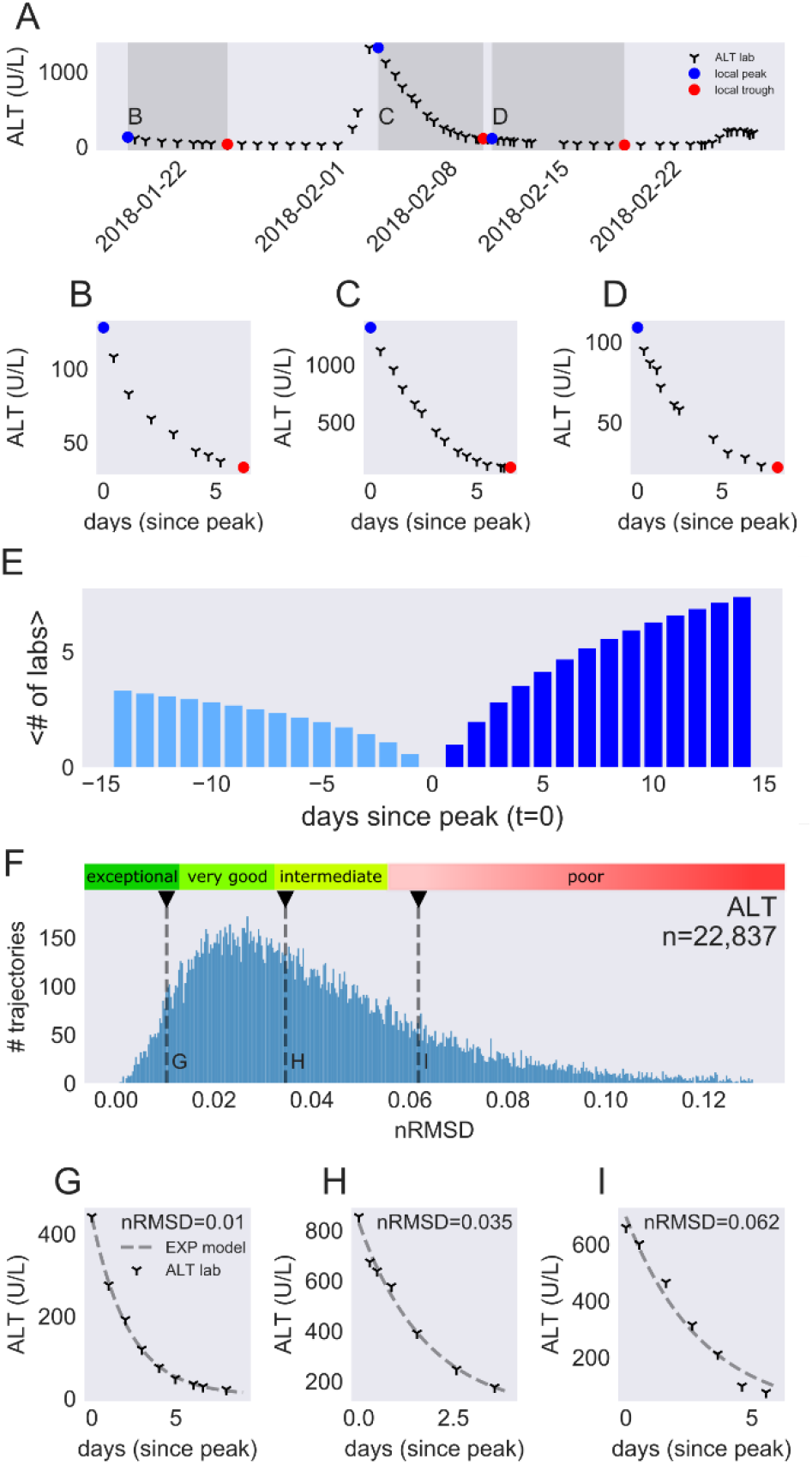
Signal-processing for ALT-trajectories. (**A**) A one month sub-interval of a patient’s full ALT trajectory, which were annotated for peaks (blue dots) and troughs (red dots). Dark gray zones correspond to (**B-D**) showing three declining trajectories from A. These trajectories are the data that model fitting operates on. (**E**) Cumulative mean number of ALT lab draws at increasing distance before or after an annotated peak. (**F**) Distribution of goodness-of-fit in nRMSD for every declining ALT trajectory meeting criteria in our dataset. Marked on the distribution are representative fits shown in sub-panels with an exceptional fit (**G**), an intermediate fit (**H**), and a poorly fit (**I**) trajectory.

The simplest model of ALT or AST dynamics can be encoded in the following differential equation:

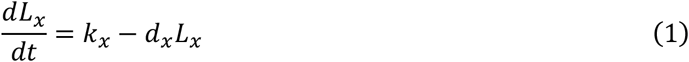

In Eq 1., the rate of transaminase (*x=*AST or ALT) value change (*L*_*x*_) over time (*t*) is proportional to the rate of hepatocyte death as measured by spillage of transaminase into the blood *k*_*x*_ (IU·L^-1^·days^-1^) minus the disappearance of enzyme at rate *d*_*x*_ (days^-1^) times the amount of enzyme present. This simple equation yields the following solution after solving the initial value problem:

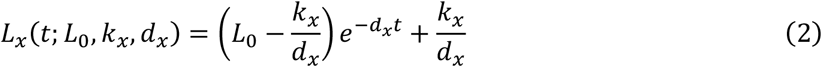

Here, *L*_*0*_ is the initial transaminase value at time zero, defined as the peak of a transaminase enzyme trajectory.

The *lmfit* python package *(15)* and Eq. 2 were used to fit the model against every trajectory meeting the following criteria: (1) at least five ALT or (separately) AST values from peak to trough, (2) monotonically declining observations, and (3) a peak value of at least five times the upper limit of normal (150 IU/L) to indicate a clinically significant event of liver injury. These criteria yielded 22,837 ALT trajectories among 20,506 unique patients, and 15,751 AST trajectories among 14,277 unique patients. Three sample ALT trajectories from the same patient (Fig. 1A) are illustrated in Fig. 1B-D. For each AST and ALT trajectory we fit the model for estimates of the parameters *k*_*x*_ and *d*_*x*_ and scored each model’s goodness of fit using the normalized root-mean-square deviation (nRMSD) (ALT-Fig 1F, AST-Fig S1; see Materials and Methods, “Model fitting**”**).

Trajectories with model fits of nRMSD<.015 showed nearly imperceptible differences between model and data (Fig. 1G, also Fig. S2-ALT, Fig. S3-AST), while models with nRMSD values as high as ∼.03 demonstrated minimal to subtle deviation from the model (Fig. 1H). nRMSD fits greater than .05 revealed poor fits (Fig. 1I). We label these categories as exceptional (nRMSD<.015), very good (nRMSD<.03), intermediate (nRMSD<.05) and poor (nRMSD≥.05). By these thresholds, 10.8% of the ALT curves were exceptional, and 40.1% were very good. Similar estimates were obtained for AST curves, with 13.2% being exceptional and 37.5% being very good. Although intermediate curves often fit the data well (Fig. 1H), we tended to use only exceptional or very good fits in our analyses as a conservative measure.

These data strongly indicate that the first-order decay kinetics encoded by Eq. 1 are seen in more than a third of clinical recovery trajectories. The presence of a first-order kinetic process reveals that a single rate-limiting biochemical step is responsible for disappearance of enzyme from the blood in these patient encounters.

### Plasma clearance is the rate limiting step in first-order kinetic disappearance of transaminase activity

At least two biochemical steps are necessary for a clinically observed downtrend of transaminase activity. First, the liver injury process (hepatocyte death) must decline or cease, thereby reducing the spillage of transaminase protein into the blood. Second, the serum AST and ALT proteins must be cleared from the blood.

Mathematical modeling of this sequence of hepatocyte death followed by enzyme clearance reveals non-first order kinetics (Materials and Methods, Eq. 8), however, two hypothetical scenarios could transform this result into first-order kinetics. One possibility is that liver injury could resolve slowly while AST and ALT enzymes are cleared from the blood quickly. The rate-limiting step in this scenario would be hepatocyte death, while the AST and ALT are instantaneously in equilibrium with the hepatocyte death rate *d*_*H*_ (Fig. 2A,B). This model reflects the traditional assumption of AST and ALT interpretation in clinical medicine where AST and ALT levels proportionally reflect hepatocyte death *(1, 4, 5)*. Alternatively, the rate limiting step is AST and/or ALT clearance from the blood, with liver injury (*d*_*H*_) rapidly resolving while AST and ALT recovery lag behind (Fig. 2C,D). Though liver injury and transaminase clearance could share a similar rate (Fig. S4E,F), this scenario would not result in a single-exponential curve, and therefore does not apply to ∼40% of trajectories defined by a single-exponential.

**Fig. 2.**
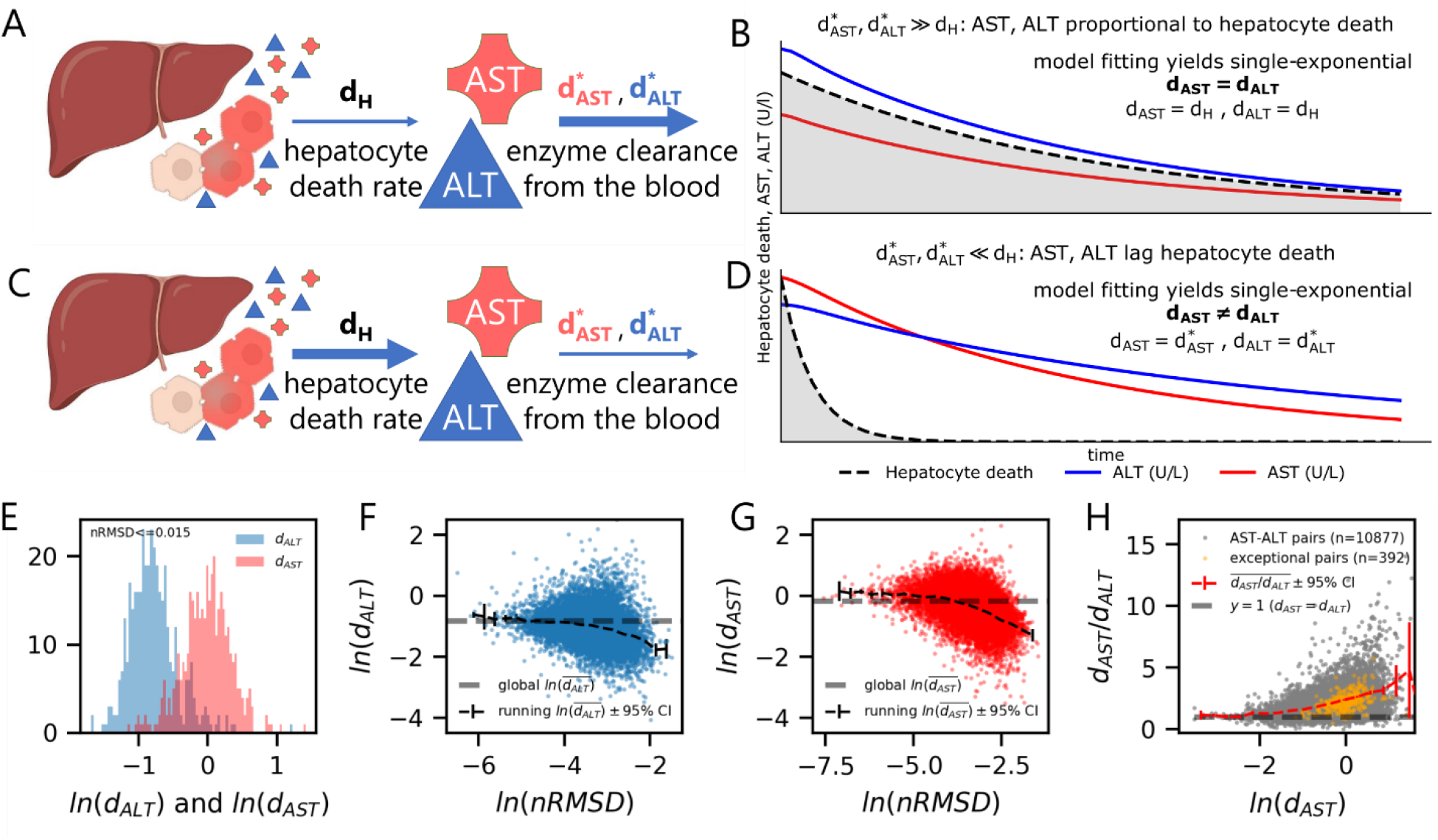
Competing mechanisms for the single rate-limiting step behavior in declining transaminase trajectories. In (**A-D**), competing hypotheses for the mechanism of a single rate-limiting step are simulated. In (**A**), hepatocyte death resolution (*d*_*H*_) is rate-limiting, while the plasma clearance rates (*d*^*\**^_x_) of each transaminase is fast. (**B)** This results in a scenario where AST and ALT are proportional to each other and hepatocyte death. (**C**) If instead enzyme clearance from the blood is rate-limiting, hepatocyte death (gray, **D**) falls off rapidly while AST and ALT decline slowly and at independent rates. (**E**) Pairs of fitted *d*_*AST*_ and *d*_*ALT*_ values from the same clinical episode where trajectories are exceptionally well-fit (nRMSD<.015) are plotted as a histogram, and show visibly different distributions. Fitted results for *d*_*ALT*_ (**F**) and *d*_*AST*_ (**G**) are plotted against their goodness-of-fit, demonstrating that as goodness-of-fit increases (farther left), a narrower estimate of the apparent clearance rate is obtained. Black lines indicate a running average, while the gray line is bulk average. In (**H**), the ratio of *d*_*AST*_ to *d*_*ALT*_ is plotted versus *d*_*AST*_, demonstrating that the ratio (running average, red) asymptotically approaches unity (dashed line, gray) where *d*_*AST*_ *= d*_*ALT*_. In contrast, the bulk of the trajectories pairs occur at higher values of *d*_*AST*_ where *d*_*AST*_ *≠ d*_*ALT*_.

These modeling predictions yield a simple experiment that can distinguish point to whether hepatocyte-death or enzyme clearance is the rate-limiting step. If hepatocyte death is the rate-limiting step, AST and ALT should decline at the same rate during the same clinical episode, as both enzymes’ trajectories are tethered to underlying liver injury recovery. That is, the model-fitted *d*_*AST*_ should be equal to *d*_*ALT*_, where both equal the hidden hepatocyte death rate *d*_*H*_ (Fig. 2B). In contrast, if liver injury rapidly recovers and AST and ALT clearance from the blood is rate-limiting, the fitted values of AST (*d*_*AST*_) and ALT (*d*_*ALT*_) should be different, as they are now untethered to hepatocyte death (see Materials and Methods, “Candidate kinetic models of transaminase behavior”). In this scenario, the fitted values *d*_*AST*_ and *d*_*ALT*_ would approximate the plasma clearance rates *d*^*\**^_*AST*_ and *d*^*\**^_*ALT*_ respectively (Fig. 2D). In enzyme kinetics, the log of the decay plot is often taken because a line in log-space corresponds to a first-order kinetic process, and the slope of that line is equal to the decay constant (Figs. S4B,D,F) *(16)*. When hepatocyte death is rate-limiting (Fig. S4B), the slopes of AST, ALT, and the hidden hepatocyte death rate are same, while all three slopes are different if enzyme clearance from the blood is rate limiting (Fig. S4D). This straightforward prediction of either the same, or different, estimates of AST and ALT *d*_*x*_ values within a single clinical encounter provides an opportunity to assess which scenario predominates clinically.

To operationalize this experiment, we first identified 10,877 trajectory pairs where the model returned a fit for both AST and ALT during a single patient encounter with liver injury. We then limited our analysis to exceptional fits (n=392) of the single first-order kinetic model (Eq. 2) to rigorously exclude the scenario where *d*_*H*_, *d*_*AST*_, and *d*_*ALT*_ are of similar magnitude (Fig. S4F). Among these unequivocally first-order kinetic clinical episodes, the fitted parameters for matched AST and ALT trajectories were significantly different with mean *d*_*AST*_ of 1.05+/-0.013 days^-1^ (SEM) and mean *d*_*ALT*_ of 0.48+/-0.008 days^-1^ (two-sided paired t-test, p=4.15×10^−280^, Fig. 2E). Fitted *d*_*AST*_ and *d*_*ALT*_ are plotted for all 10,877 trajectories against goodness-of-fit (nRMSD) demonstrating that better fit values tend towards the scenario where *d*_*AST*_ ≠ *d*_*ALT*_. This indicates that among clinical episodes well-modeled by first-order kinetics, plasma clearance of transaminases is rate-limiting. The clinical implication is that observed AST and ALT levels considerably lag behind the cessation of liver injury (Fig. 2C,D).

Although the dominant pattern among well-fit curves supports the model where AST and ALT clearance are rate-limiting, we hypothesized that there should exist a clinical episode in our large dataset where liver injury recovery is slow enough to be rate-limiting. To seek out these scenarios, we plotted *d*_*AST*_ against the ratio of *d*_*AST*_/*d*_*ALT*_ for all paired AST:ALT trajectories. We observed that in clinical encounters with very low fitted *d*_*AST*_ values the ratio converges on 1 (Fig. 2M, dashed line), capturing the expected situation for when *d*_*AST*_ = *d*_*ALT*_ = *d*_*H*_. This supports the scenario in Fig. 2A-C, which should only be observed when *d*_*H*_ is sufficiently slower than *d*^*\**^_*ALT*_ and *d*^*\**^_*AST*_. Finally, it is notable that although hepatocyte death can be the single-rate limiting step in our dataset, it constitutes only 0.3% of all matched AST:ALT trajectories (Fig. 2H and Supplementary Materials, “Hepatocyte death is rarely rate-limiting among first-order kinetic trajectories”).

To summarize, transaminase recovery trajectories that are well-fit by first-order kinetics demonstrate different estimates for *d*_*AST*_ and *d*_*ALT*_. This strongly supports the hypothesis depicted in Figs. 2C and 2D where declining transaminase levels reflect clearance of AST and ALT from the blood, rather than cessation of liver injury. Clinically, this means improvement of measured AST and ALT values significantly lags cessation of hepatocyte injury. In the competing hypothetical scenario (Fig 2A,B) where hepatocyte death is rate-limiting, clinical observations validate our modeling prediction that *d*_*AST*_ = *d*_*ALT*_, where both values are equal to the hidden hepatocyte death rate. However, this scenario is extremely rare (left-most aspect of Fig 2H).

### Estimation of the plasma clearance rates for AST and ALT in humans

A major result of the above analysis is that instead of representing the dynamics of liver injury resolution, transaminase recovery trajectories frequently reflect the plasma clearance of AST and ALT. While many scenarios modeled likely exemplify some mixture of hepatic injury recovery and plasma clearance, progressive filtering of goodness-of-fit should result in fitted *d*_*AST*_ and *d*_*ALT*_ converging on plasma clearance rates of AST (*d*^*\**^_*AST*_) and ALT (*d*^*\**^_*ALT*_) (Fig. 2C). This is precisely what is seen in Figs. 2F and 2G; as goodness-of-fit improves (smaller nRMSD), the fitted values of *d*_*AST*_ and *d*_*ALT*_ asymptotically converge on the putative plasma clearance rates of AST (*d*^*\**^_*AST*_) and ALT (*d*^*\**^_*ALT*_). The binned average indicates the mean value obtained by examining only exceptional fits is an accurate representation of *d*^*\**^_*ALT*_ and mildly underestimates *d*^*\**^_*AST*_. Variance minimization is a more objective way of choosing nRMSD cutoffs that assure first-order kinetics and yielded similar predictions: *d*^*\**^_*AST*_=1.13d^-1^ and *d*^*\**^_*ALT*_=0.47d^-1^ (Fig. S5) (see Supplementary Materials, “Variance minimization to estimate the plasma clearance rates of AST and ALT”). These results reveal AST to be cleared from the blood at more than twice the rate than ALT.

To investigate the mechanism for plasma clearance, we examined whether the transaminase plasma clearance rates changed with kidney or liver injury, the two primary organs responsible for removing proteins from the blood *(17)*. We considered peak transaminases to reflect the degree of liver injury, PT-INR to reflect the degree of liver dysfunction, and total bilirubin to reflect both injury and dysfunction. Creatinine levels were used to model kidney injury and dysfunction. For all AST and ALT trajectories meeting fitting criteria, we correlated PT-INR, total bilirubin, peak transaminase and creatinine with model-fit *d*_*x*_. To enrich for clinical scenarios that were more representative of plasma clearance, we initially limited our analysis to trajectories with very good or exceptional fits (nRMSD<.03). For ALT trajectories with a model-fitted *d*_*ALT*_ (n=9,173), there was no correlation with peak ALT (Pearson’s correlation, r=.015), PT-INR (r=-.002), total bilirubin (r=.026), or creatinine (r=.020) (Supplementary Table S1). Similarly, *d*_*AST*_ (n=5,917) was not correlated with peak AST (r=.067), PT-INR (r=-.012), total bilirubin (r=-.022), or creatinine (r=-.023) (Supplementary Table S2). Relaxing the requirement for reasonable goodness-of-fit did not substantially change the correlations (Supplementary tables S3 and S4). To further enrich for scenarios where organ failure is likely to influence plasma clearance rate we filtered for patients with renal failure (Cr>2.0) and liver failure (INR>2, TBili>3), however, these subgroups also did not demonstrate significant correlation with plasma clearance rate (Supplementary Tables S5-8).

Gender and age-specific changes in the plasma clearance rate were also examined. Age did not correlate with plasma clearance rate for AST (r=-0.01) or ALT (r=-0.01). In contrast, clearance rates for both AST and ALT for males (d^*^_AST_=0.45d^-1^, d^*^_ALT_=0.93d^-1^) was significantly albeit modestly lower than for females (d^*^_AST_=0.48d^-1^, d^*^_ALT_=1.03d^-1^; t-test, d^*^_AST,_ p= 1.04×10^−11^, d^*^_ALT,_ p= 4.2×10^−5^).

These data suggest (1) mathematical modeling allows inference of the plasma clearance rates of AST and ALT in humans, (2) the plasma clearance rates are not influenced by liver or kidney dysfunction and (3) that the plasma clearance rates are unaffected by age but are ∼10% faster in females compared to males.

### Transaminase kinetic behavior distinguishes active hepatitis from normal histology

Estimation of the plasma clearance leads to a testable prediction about whether liver injury is active or resolving during individual episodes of liver injury recovery regardless of absolute transaminase level. When a declining trajectory’s fitted *d*_*x*_ is similar to the plasma clearance rate *d*^*\**^_*x*_, we predict that liver injury is resolving or has since resolved, as depicted in Fig. 2C. In contrast, fitted *d*_*x*_ values progressively lower than the plasma clearance rate *d*^*\**^_*x*_ correspond to increasingly active hepatocyte death, despite enzyme downtrend.

To test this prediction, we examined individual patients in the identified cohort with liver biopsy results during a model-fitted ALT and AST trajectory. We identified 141 datasets comprising a trio containing (1) a fitted AST trajectory, (2) a fitted ALT-trajectory, and (3) a matched liver biopsy occurring during downtrend of a trajectory before AST and ALT normalized (Data S1). For each pathology report, the final diagnosis was scored for a “dominant process” (see Supplementary Materials, “Scoring liver histopathology reports for a dominant process”). The most common dominant process was “hepatitis” (n=35) which we compared to cases documented as “normal” (n=11). There was a marked reduction in the fitted *d*_*x*_ values for “hepatitis” (*d*_*AST*_*=*0.40±.048h^-1^, *d*_*ALT*_ =0.21±.03h^-1^) as compared to “normal” (*d*_*AST*_*=*1.05±0.13h^-1^, *d*_*ALT*_ =0.51±.21h^-1^) (t-test for both, p<.0001, see Figs. 3A and 3B). The mean *d*_*AST*_ (Fig. 3A) and *d*_*ALT*_ (Fig. 3B) for normal cases was similar to the estimated plasma clearance rate. This could not be entirely explained by lower absolute transaminase values among cases labeled as “normal” (AST=485±289 IU/L, ALT= 362±115 IU/L 95% CI) compared to “hepatitis” (AST=1025±339 IU/L, ALT=1116±406 IU/L, 95% CI) as normal cases still fell into the category of severe liver injury on average (>10 times the upper limit of normal) *(18)* and ranged as high as AST=1331 IU/L and ALT=768 IU/L (Fig. 3C, 3D). Further acknowledging portal hepatitis is considered less severe than lobular hepatitis, and that cholestasis is considered a signature of healing after injury, a comparison of mean *d*_*AST*_ and *d*_*ALT*_ values shows a clear ordering which matches expectation (lobular hepatitis<portal hepatitis<cholestasis<normal) (Fig. S8, Table S8).

**Figure 3.**
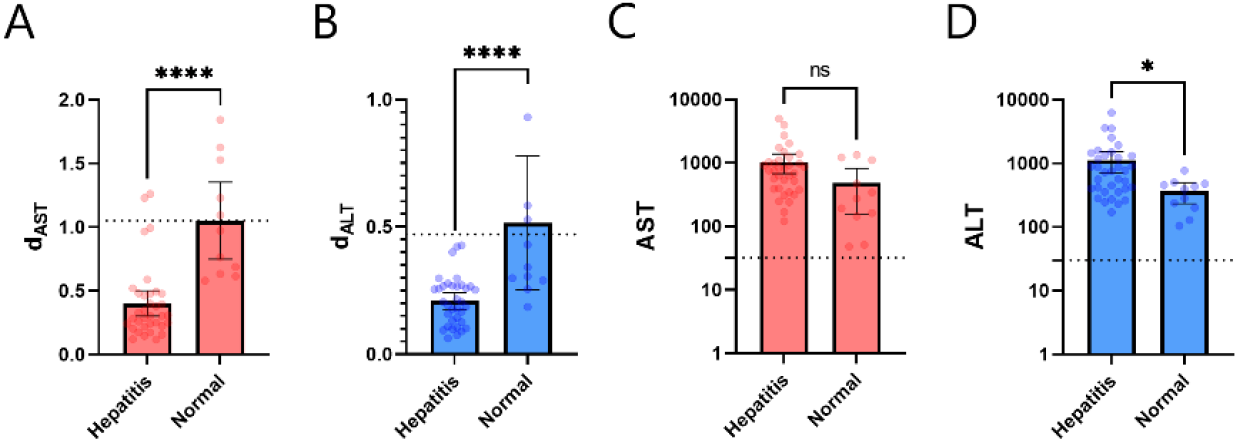
Kinetic decline in AST and ALT in first-order trajectories differentiates hepatitis from normal pathology in contemporaneous liver biopsies. (**A** and **B**) Fitted *d*_*AST*_ and *d*_*ALT*_ are markedly lower in trajectories where pathology showed active lobular inflammation (hepatitis) compared to trajectories where pathology showed normal liver parenchyma (p<.0001). Plasma clearance rates *d*^*\**^_*AST*_ and *d*^*\**^_*ALT*_ are shown by dotted lines. (**C** and **D**) Nearest AST or ALT values to the time of liver biopsy were not significantly different (AST, **C**, p=.010) or marginally lower (ALT, **D**, p=.049) in cases of active hepatitis versus normal liver parenchyma. The upper limits of normal for AST and ALT are marked by the dotted lines.

These results indicate that in the case of active hepatitis where the pathologic description of ongoing injury is unambiguous compared to normal histology, the rate of AST and ALT decline as summarized by the model-fitted *d*_*AST*_ and *d*_*ALT*_ parameters strongly distinguishes between ongoing liver injury and resolution. The intermediate histopathology findings of portal hepatitis and cholestasis have *d*_*AST*_ and *d*_*ALT*_ parameters that fall in between lobular hepatitis and normal. Cases documented as normal converge on estimates of the plasma clearance rate. This analysis provides histopathologic evidence that first-order kinetic decline reflects clearance of enzyme rather than hepatic injury (Fig. 2D) and validates our estimates for the plasma clearance rates of AST and ALT in humans.

### Derivation of a real-time hepatic injury index

Modeling coupled with clinical validation above demonstrates that the kinetics of clearance versus injury are captured by a fitted parameter in trajectories with sufficient observations. While informative for retrospective data as we have shown, such an approach has limited potential for prospective clinical application given the need for five or more observations and a sophisticated fitting routine. To better estimate real-time hepatic injury with fewer observations, we directly incorporated prior knowledge of the plasma clearance rates for AST and ALT and derived a Hepatic Injury indeX (HIX) which can be calculated from any pair of AST or ALT observations (Eq. 3—see Materials and Methods, “Derivation of the HIX”).

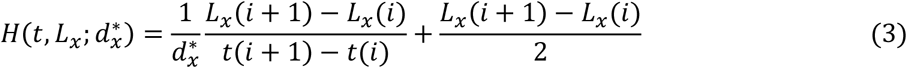

The HIX index, H(t) was derived from the following intuition. For a fixed plasma clearance rate which we estimated above, and any given transaminase value, the precise trajectory transaminases should follow if liver injury were to resolve can be directly calculated. Any deviation from this complete resolution trajectory must be due to ongoing liver injury; the index calculates a specific representation of that difference which mimics conventional interpretation of AST and ALT. In Fig. 4A, the derivation of the HIX is shown graphically; for any two ALT values coupled with the known plasma clearance rate of the enzyme, a trajectory can be drawn through the values which levels off at some later time. That leveling off point is the level of ongoing injury *after enzyme clearance from the blood equilibrates to the degree of ongoing hepatocyte death*. In Fig. 4A, the second ALT value is above the expected resolution trajectory if liver injury had ceased at time zero, therefore, despite downtrend, liver injury is ongoing. The HIX predicts that if the degree of liver injury remains the same, the ALT values 6 days later will level off at AST=1100 IU/L. Therefore, the HIX value is 1100 IU/L.

**Figure 4.**
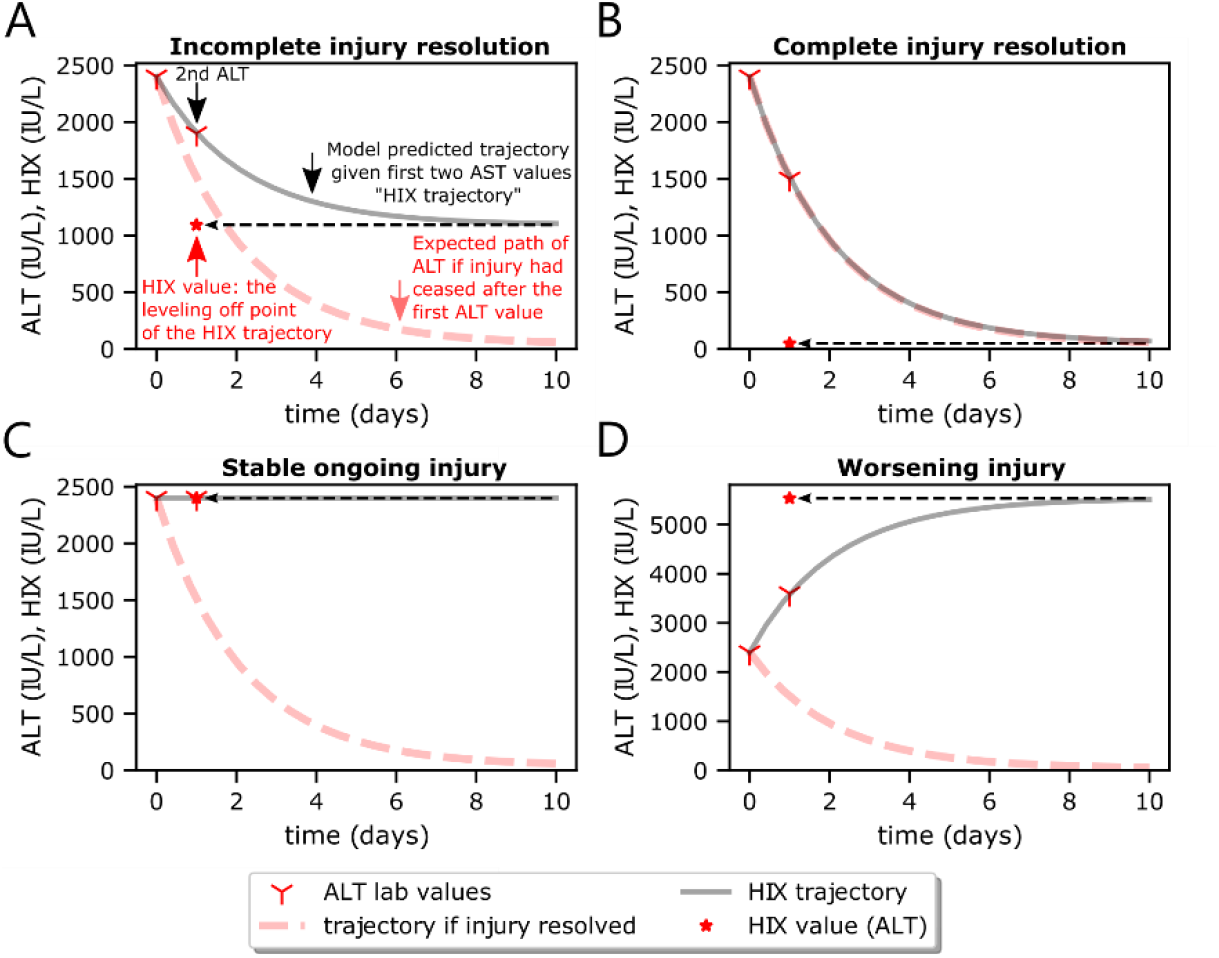
The Hepatic Injury indeX (HIX) corrects for lag in ALT due to plasma clearance. In (**A-D**), two serum ALT values are shown (red markers) in simulation. Assuming liver injury resolves on day 0, the anticipated trajectory from the first ALT value is expected to follow kinetics dictated by plasma clearance alone (red dashed line). In (**A**), the second ALT is above the complete recovery curve indicating a degree of ongoing injury. Accounting for plasma clearance (gray line), the level of ongoing liver injury in units of ALT is equal to the leveling off point of the HIX trajectory, and yields the HIX value (asterisk). In contrast (**B**), when the second value falls on the predicted plasma clearance trajectory, the HIX trajectory is equivalent to plasma clearance indicating cessation of liver injury, and results in HIX=18 IU/L. Whether liver injury has resolved completely (B), is stably elevated (**C**), or is worsening (**D**), the HIX reflects the real-time level of liver injury in units of ALT (IU/L) after accounting for plasma clearance.

When liver injury has resolved, the second ALT value will follow the kinetic trajectory of enzyme clearance alone, and therefore falls directly on the injury resolution trajectory (Fig. 4B). In this case, the HIX=18 IU/L on day 1, reflecting that even with an absolute level of ALT=1500 IU/L, the ALT is only elevated because enzyme has not been cleared from the blood, while liver injury has resolved. When liver injury is stably elevated between two sALT values (Fig. 4C), there is a biophysical steady state between plasma clearance and production of ALT due to liver injury, therefore the ALT=HIX=2400 IU/L. The HIX is calculable for rising trajectories as well and again predicts the leveling off point of ALT assuming the degree of liver injury remains the same (Fig. 4D).

In summary, the HIX corrects for the delay inherent to ALT and AST due to slow clearance of enzyme from the blood. The HIX does not make predictions about liver injury per se, but clarifies more precisely what AST and ALT would otherwise report if their half-lives were much shorter. The HIX value can be calculated with each new laboratory draw of AST or ALT and can guide clinical decision-making 3 days (AST) or 6 days (ALT) ahead of usual clinical practice of trending transaminases to their peak or baseline.

### The HIX predicts histological inflammation and injury resolution independent of absolute AST and ALT levels

To understand whether the HIX makes useful clinical predictions we again examined cases where liver biopsies were undertaken in close proximity to laboratory measurements of transaminases where the HIX could be calculated. This yielded 810 pairs of HIX assessments with liver histopathology, of which 621 were confirmed to be parenchymal core needle biopsies of the liver (Data S2). Liver histopathology was scored for a dominant process. The resulting dataset included a label for histopathology, the nearest HIX value calculated from the AST or ALT values flanking the biopsy date and time, and the nearest AST and ALT value. Of particular interest were scenarios where the absolute transaminase value deviated the most from the HIX value, which was quantified by calculating the relative fractional change of the HIX in comparison to the AST or ALT (Figs. 5A,B). Thus, negative values indicate the HIX predicts liver injury to be less than the AST or ALT, while positive values indicate the HIX predicts liver injury to be more severe than the AST or ALT. Liver biopsies revealing normal histology were significantly biased toward having HIX values much less than AST (Fig. 5A, p<.0001) and ALT (Fig. 5B, p=0.048). In contrast, lower absolute transaminase values cannot explain differences in pathology for AST (Fig. 5C, p=0.71) or ALT (Fig. 5D, p=0.33). Examination of individual patient trajectories highlights cases where the HIX warns of active injury despite a downtrend (Fig. 5E), mirroring the simulation in Fig. 4A, and cases where injury has essentially resolved despite high absolute levels of AST and ALT (Fig. 5F), mirroring the simulation in Fig. 4B. These cases share nearly the same absolute ALT value at the time of biopsy (ALT=1285, Fig. 5E; ALT=1290, Fig. 5F). These data indicate that that the HIX more accurately depicts a real-time assessment of liver injury than the absolute level of AST or ALT.

**Figure 5.**
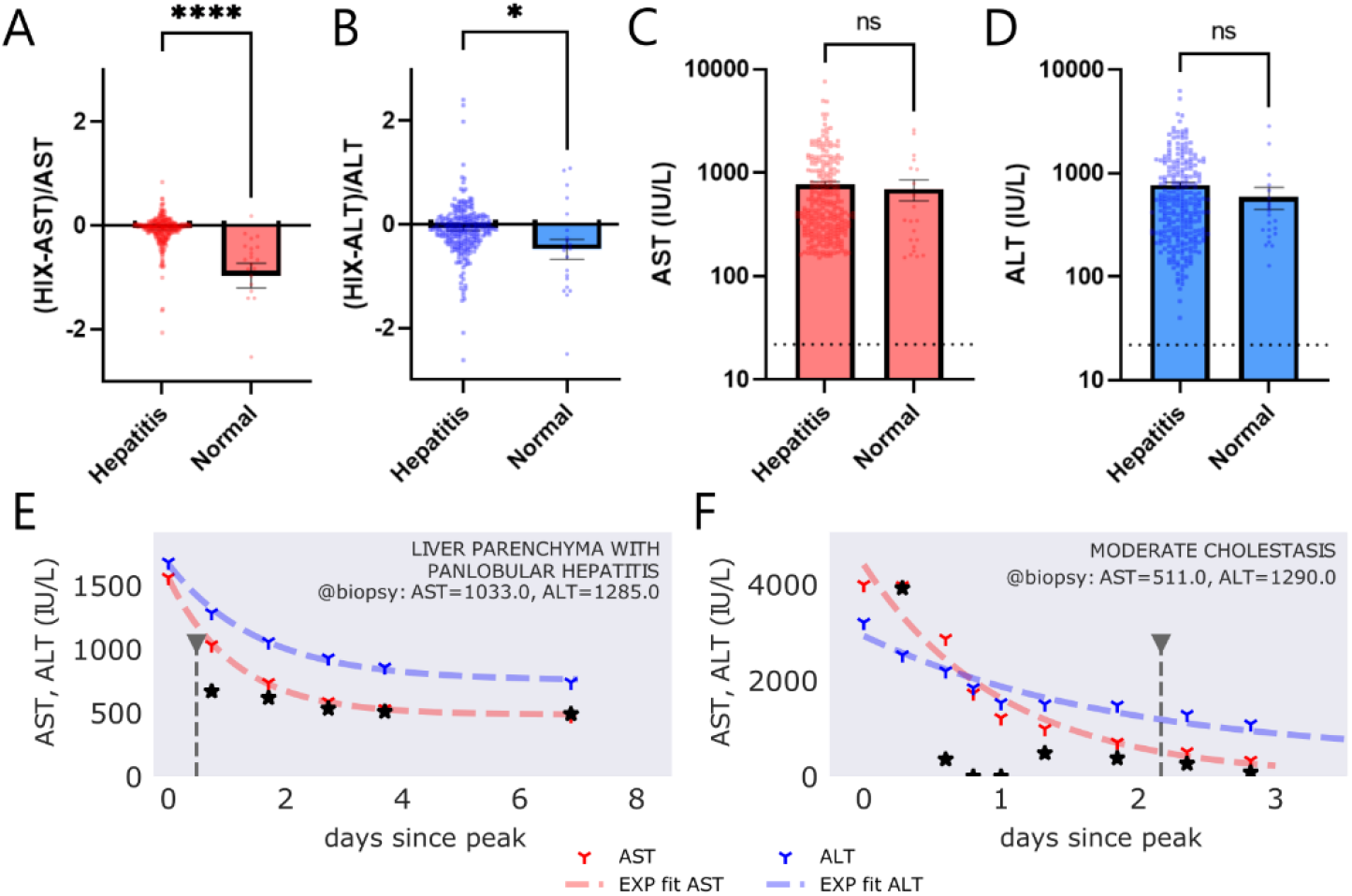
The Hepatic Injury indeX (HIX) predicts histologic inflammation and resolution independent of absolute transaminase levels. Relative difference between the HIX and AST (**A**) or ALT (**B**) closest to the point of a liver biopsy are compared between biopsies showing lobular hepatitis (n=253) versus normal liver parenchyma (n=20). Absolute AST (**C**) and ALT (**D**) closest to the time of liver biopsies showing hepatitis or normal pathology. (**E and F**) Two patient AST and ALT trajectories with the same absolute ALT value at time of biopsy are plotted along with model fit, HIX prediction, and the timing of the liver biopsy during the encounter. (**E**) AST and ALT are in downtrend, however, the HIX anticipates leveling off of AST (and therefore active liver injury) 3 days prior to AST and ALT leveling off; biopsy confirms active hepatitis during the downtrend. In contrast, (**F**) AST and ALT are in downtrend but the HIX predicts resolution of liver injury despite ALT=1290, AST=511 at time of biopsy. The biopsy confirms evidence of recent but no ongoing injury.

To further test the utility of the HIX, specific scenarios were identified where the HIX result differed the most from standard clinical interpretation. We first sought patient encounters where the nearest AST was > 150 IU/L and where the HIX predicts injury resolution in that interval (HIX<35 IU/L). This yielded 17 clinical scenarios with associated liver histopathology (Table S10). For each of these 17 patients where the model predicts complete resolution of injury we selected a control matched on absolute AST, with identical criteria, except that the HIX was predicted to be > 40 IU/L. The resulting 17 cases with nearly identical AST values represent case pairs that are clinically comparable by current standards. Pathology reports were manually scored for normalcy, degree of lobular inflammation, degree of portal or central inflammation, and degree of necrosis in a blinded fashion, with results shown in Table S10. Among these 17 examples, 17/17 (100%) showed mild inflammation or less in the HIX-resolved category, with 5/17 (29.4%) being read as normal. In comparison, none of the matched controls had normal pathology (p=.022, Fisher’s exact), and paired comparisons of lobular injury (p=0.007) and portal/central injury (p=0.024) were significantly greater in the high HIX cohort (paired Wilcoxon signed-rank test). The degree of necrosis was not statistically significantly different, with both categories showing a large spectrum of necrosis degree (p=0.69, Wilcoxon signed-rank test). Illustration of specific examples from Table S10 with matched controls are shown in Fig. S7.

These findings demonstrate that the HIX is deployable to any pair of AST or ALT measurements and that the HIX reliably distinguishes normal to mild inflammation from moderate to severe inflammation independent of absolute AST or ALT levels. The interpretation of the HIX exactly replaces the way AST or ALT is currently interpreted in clinical practice: the HIX is proportional to the degree of ongoing liver injury, and it is reported in units of AST or ALT.

## Discussion

We analyzed 6.5 million individual AST and ALT measurements and 90,770 patients’ transaminase trajectories using database programming and mathematical modeling to estimate the plasma clearance rate of AST and ALT in humans. A substantial proportion of trajectories can be described by first-order kinetics, indicating a single rate constant determines their kinetic behavior. We demonstrate that it is the elimination of transaminase activity from plasma, rather than ongoing liver injury, that results in first-order kinetics. This discovery enabled us to derive a correction to AST and ALT, the HIX, which removes the effect of enzyme clearance, thereby providing a more faithful real-time estimate of liver injury. As proof of concept, we identified liver biopsies obtained during dynamic change in AST and ALT where the HIX and the absolute levels of AST and ALT were maximally discrepant. We determine that the HIX can identify resolution of liver injury with accompanying unremarkable liver histology despite marked persistent elevation of transaminase, demonstrating that the HIX more sharply defines presence or absence of liver injury.

A major finding of this study is an estimate of the *in vivo* plasma clearance rates for AST and ALT in humans. Traditionally, clearance rates have been obtained by *in vivo* approaches through direct injection of a labeled enzyme into a model organism with no ongoing liver injury and tracking decline in serum enzyme over time. This has been done with healthy pigs (AST t_1/2_=18h, ALT t_1/2_=51h), dogs (ALT t_1/2_=45.2h), and rats (AST t_1/2_=17.8h) among others *(16, 19–22)* (Table S11). These half-lives are in good agreement with our human estimates (AST t_1/2_=15.8h, ALT t_1/2_=34.6h). Curiously, clearance rates are often cited as known quantities in humans *(4, 18, 23–29)*, even though the definitive study of monitoring enzymes’ decay in healthy human subjects has not been undertaken. Rather, the values for these apparent half-lives originate from only three original sources in the literature, where the majority of references point to reviews by Price and Alberti *(27)* or Schiff *(26)* who cite human half-lives of AST t_1/2_=17h and ALT t_1/2_=47h without providing a direct reference. These numbers, however, are precisely the half-lives estimated in a German-language manuscript by Bär and Ohlendorf *(30)* from 1970 based on declining AST or ALT trajectories in six patients observed with enzyme activity determined every 12 hours for 72 hours after acute myocardial infarction (4 patients), myoglobinuria (1 patient) or liver surgery for echinococcal cysts (1 patient). Two additional sources examined ALT decline after hepatitis (n=5, ALT t_1/2_=48h) *(31)* or after acute myocardial infarction (n=40, AST t_1/2_=20h) *(25)*. An essential difference of our study, besides a vastly increased number of patients, is that prior estimates of AST and ALT clearance assume liver (or heart) injury to have ceased immediately at the onset of enzyme downtrend. Here, we avoid this assumption by taking advantage of an extraordinary number of patients to filter for trajectories that were highly first-order in character, as ongoing injury with influx of new enzyme is expected to cause trajectories to deviate from first-order kinetics. The presence of modest ongoing injury in prior studies likely explains why we saw shorter half-lives for both AST and ALT. Further, we recognized that different biomarkers of the same biological process allow distinguishing between ongoing organ injury and plasma clearance (see Fig. 2A-C). These two principles offer a powerful complementary approach to traditional model organism-based biochemical estimation of enzyme plasma clearance, while vastly increasing the potential utility of widely available electronic medical records.

Our massive dataset allowed us to estimate clearance rates across age, gender, and concomitant disease and thereby glean insight into the process by which AST and ALT are cleared from the blood. Severe kidney disease did not significantly affect clearance of enzyme, which is consistent with the size restriction of glomeruli *(32)* (<60 kda) for AST and ALT dimers (94 kDa *(33)* and 110 kDa *(34)*, respectively). Interestingly, severe hepatic dysfunction also did not significantly alter the estimated clearance rate. While loss of enzyme activity or proteolytic degradation in circulating plasma is possible, elegant studies in rats offer a more compelling mechanism *(16, 20)*: injection of radiolabeled AST localizes to hepatic endothelial cells, but not hepatocytes. In these animals, 90% hepatectomy yielded marked retardation of plasma clearance, indicating the liver is the primary source of degradation. Limited 50% hepatectomy, however, had no effect on enzyme clearance indicating a large capacity for clearance *(20)*. If hepatic endothelial cells are the source of degradation in humans, we speculate that sufficient liver endothelial function exists even in liver failure to preserve enzyme clearance. More direct experimental efforts would be required to better establish the mechanism of AST and ALT clearance in humans.

This stability of enzyme clearance rates across demographics and disease states allowed us to derive a universal hepatic injury index (HIX), an equation that estimates *bona fide* liver injury from any two sequential measurements of AST or ALT. One may think of any AST or ALT value as being the sum of two components: transaminase produced at that moment from ongoing liver injury, and transaminase produced from prior injury that has not yet been cleared from the serum. The functional difference between the HIX and the AST and ALT is shown in Fig. 6. If liver injury is constant, and maximal at time 0, the expected pattern of AST and ALT laboratory values are shown in simulation using the kinetic rate constants derived in this study. The corresponding HIX values calculated from the AST and ALT values are also shown and demonstrate the HIX provides instantaneous estimation of the true hepatocyte death rate, while it would take 4 half-lives (AST = 2.6 days; ALT = 5.8 days) for transaminases to reflect the actual level of liver injury. Our work highlights that AST and especially ALT are lagging indicators of liver injury, and that the HIX delivers a much better real-time estimate of liver injury.

**Figure 6.**
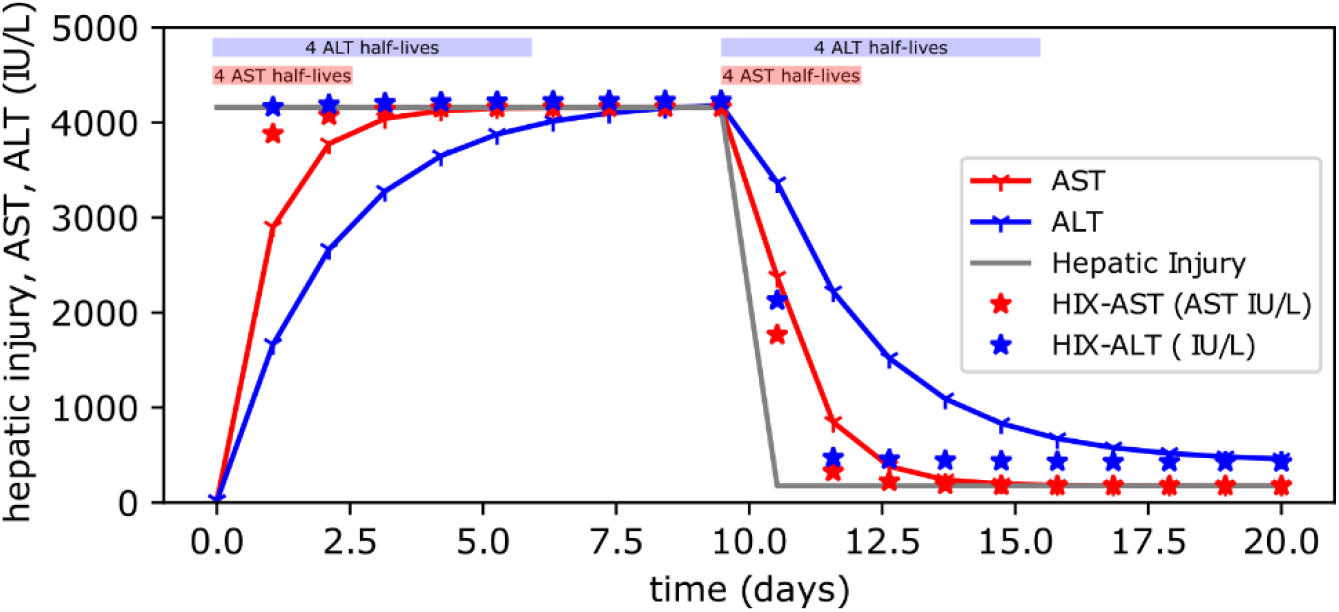
Comparison of AST and ALT biomarkers versus HIX-AST and HIX-ALT. If liver injury is maximal and constant at t=0 (gray line), and then resolves instantaneously on day 10, the expected AST (red line) and ALT (blue line) trajectories are shown. HIX values calculated on the daily labs starting at Day 1 are shown for HIX-AST (red star) and HIX-ALT (blue star). Four half-lives (93% clearance) are shown for AST (red rectangle) and ALT (blue rectangle) for both rising and falling trajectories across the top.

The HIX is an important interim achievement for improving prompt clinical decision-making, even for physicians not intimately familiar with liver disease, but ultimately only highlights the need for more responsive biomarkers. In contrast to liver disease, biomarkers for cardiac disease have been assiduously improved to more closely predict both cardiac injury onset and resolution *(35)*. One explanation for the robust innovation in the cardiac biomarker space may be that both electrocardiogram and patient symptoms preceded earliest detection of first-generation cardiac injury biomarkers (including AST), while simultaneously rapid and clinically beneficial interventions became available, highlighting a clinical need *(36)*. The fact that many liver diseases are asymptomatic up until the point of jaundice or liver failure combined with a relative lack of time-sensitive and impactful interventions may explain why AST and ALT have not been improved on since their initial description *(1, 37)*. The present study offers great impetus to improve on these biomarkers, and the potential benefit of a more responsive biomarker may not be apparent until we deploy that tool for clinical use.

In summary, this work establishes an estimate for the biophysical clearance rates of AST and ALT in humans using kinetic modeling and database programming. These observations lead to a fundamentally different interpretation of abnormal transaminases and have immediate clinical relevance. Our modeling shows how clinicians can decide, even when ALT or AST is >1000, whether an individual patient has resolved their liver injury, or when elevated concern is needed despite a downtrend in enzymes. We expect the HIX (hix.massgeneral.org) will reduce the gap where usual care is simply to monitor expectantly, leading to more timely liver biopsies when the trend is concerning, or more patient reassurance and discharges when the trend is reassuring. In addition to the possibility that such an index will sharpen clinician decision-making in the context of acute liver injury, we anticipate the approach taken here coupling database programming with mechanistic kinetic modeling will continue to produce new insights for patients with liver disease and serve as a framework for understanding other clinical time series.

## Materials and Methods

### Study Design

This retrospective, single-healthcare system, multi-center retrospective cohort study was approved as an exempt protocol by the Mass General Brigham Institutional Review Board. The aim of this study was to rigorously define the quantitative behavior of AST and ALT over time in human subjects with acute liver injury, and to use these dynamics to identify commonality and differences between patients and between disease states. The study is comprised of three components. The first was bioinformatic and descriptive, to understand the overall structure and feasibility of applying kinetic modeling to individual patient trajectories in a large database. The second component was exploratory kinetic modeling where we proposed candidate mechanistic models for the recovery phase of liver injury and fit these models to individual patient trajectories. Discoveries from this phase informed the third component, where we asked whether model results from specific transaminase trajectories predicted gold-standard liver biopsy histopathology.

### Patient data acquisition, structure, and initialization

This study comprises retrospective datasets obtained from two queries to the Partners Research Patient Data Registry (RPDR). The first dataset was a de-identified query for laboratory data comprising 91,086 (query date 1/26/21) patients with at least one ALT>150 and greater than 6 measurements of ALT. From this query we obtained all ALT and AST measurements from matching patients from 1995 to query date, along with additional laboratory results (Table S2). The resulting dataset contained 3,334,886 AST measurements, and 3,222,402 ALT measurements. For the pathology data, we made an identified query to the RPDR for patients with a liver biopsy who also had at least one measurement of ALT>=400 (query date 3/9/21). This yielded 9,274 patients, their AST and ALT laboratory measurements, and associated liver pathology reports.

For both sets of data, we performed the following pre-processing steps in Python using custom scripts. Laboratory results for transaminases were coerced to numerical form; values above or below the limit of assay were discarded. Data were assembled in a flat Pandas dataframe *(38)* with individual laboratory entries marked by categorical labels. An individual, full AST or ALT trajectory for each patient was defined as a patient’s complete laboratory history for AST or ALT, sorted by time. This was obtained by filtering the dataframe for a particular patient, as marked by an enterprise master patient index (EMPI), and the particular transaminase label (e.g., “ALT”). This full transaminase trajectory was then annotated for peaks and troughs using SciPy signal processing functions *(14)*, and peaks and troughs occurring on the edges of trajectories were discarded.

For initial model building, an individual clinical episode was defined as sub-intervals on the full patient trajectory comprising all the laboratory measurements from peak to trough. To ensure these trajectories captured a coherent, single clinical episode we screened for 5 or more consecutive, monotonically declining AST or ALT observations with the following additional stipulations. The first and second measurement were required to be less than 5 days apart. Secondly, we truncated the trajectory when the percent change between observations was <1%. This was done because once a trajectory reaches steady state, additional values occurring at steady state begin to weight goodness-of-fit for the flat part of the curve, whereas we sought goodness of fit during the dynamic portion of each trajectory. If at the end of these additional processing steps we still had 5 or more transaminase measurements, this trajectory was included.

For the matched histology analyses, pathology reports were processed using text parsing in Python to identify the “final diagnosis” text using keywords. Additional text was discarded for anonymity. Final diagnosis text was recorded in a Pandas database along with EMPI and date of biopsy, and analyses in which a biopsy occurred during a trajectory was defined simply as a biopsy date occurring within the time-range of a given trajectory. For the HIX and matched liver histopathology analyses, these requirements were relaxed to any liver biopsy for which there were two AST (or two ALT) laboratory values obtained within ±1 day of the liver biopsy date and time.

### Model fitting

For each trajectory meeting criteria, model fitting of the trajectory to Eq. 2 was attempted using the *lmfit* package *(15)* in Python using default settings. Parameters were required to be positive but were otherwise unconstrained. Model fit results were scored using normalized root-mean-square deviation (nRMSD), where RMSD was normalized by the range within each trajectory to allow comparison of fits with widely different scales:

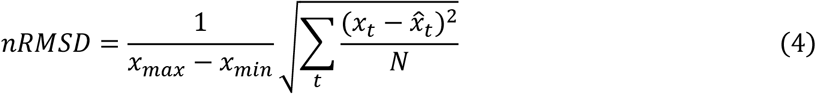

In Eq. 4, *x*_*t*_ is the lab value at time t, and 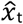 is the fitted value, thus the numerator within the summand corresponds to the residuals, and *N* is the number of laboratory values in that trajectory.

### Candidate kinetic models of transaminase behavior

Eq. 1 captures the most simplistic representation of biomarker *x*, where *x* is either AST or ALT. An important subtlety in our treatment is that *d*_*AST*_ (*d*_*S*_) and *d*_*ALT*_ (*d*_*L*_) represent the apparent degradation rate of any given curve, but there is no guarantee that any individual *d*_*x*_ reflects the plasma clearance rate *d*^*\**^_*x*_. This is shown schematically in Fig 2A-D, which represents the following system of differential equations.

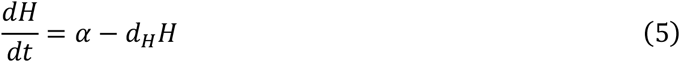

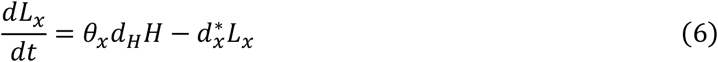

In Eq. 6, *H* is the number of hepatocytes, α reflects the hepatocyte production rate, *d*_*H*_ (days^-1^) represents the hepatocyte death rate, θ_x_ is a constant representing the transaminase concentration in blood produced per dead hepatocyte (IU·L^-1^), and *d*^*\**^_*x*_ (days^-1^) is the plasma clearance rate of *x* (AST or ALT). Eq. 5 is similar to Eq. 1, except now directly modeling hepatocyte production and death, a term previously considered to be a constant.

The simulation result from Fig. 2A-D may be shown analytically. Setting θ_x_ = 1 for convenience, the solution to Eqs. 4 and 5 are:

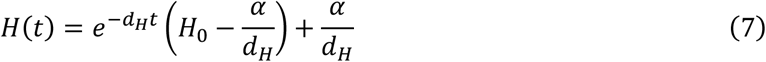

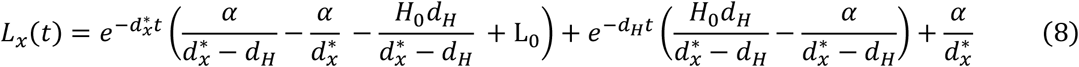

By inspection, in the limit of *d*^*\**^_*x*_ >> *d*_*H*_, the second term predominates, and the apparent decline in transaminase *L*_*x*_*(t)* is defined by single-exponential behavior with characteristic decay *d*_*H*_. This corresponds to hepatocyte death being the rate limiting step (Fig. 2A,B), with apparent *d*_*x*_ from Eq. 2 being equivalent to *d*_*H*_. In contrast, it follows that in the limit of *d*_*H*_ >> *d*^*\**^_*x*_, the second term of Eq. 7 rapidly goes to zero in comparison to the first term, and the rate of decline in observed transaminase *L*_*x*_*(t)* is dominated by a single exponential with characteristic decay *d*^*\**^_*x*_, such that Eq. 2 *d*_*x*_*= d*^*\**^_*x*_. This corresponds to Fig. 2C and D where decay over time is representative of plasma clearance.

### Derivation of the HIX

We re-ordered Eq. 1 to solve for *k*_*x*_, which represents the rate of liver injury:

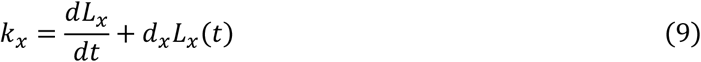

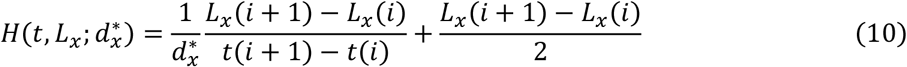

We then discretize the derivative and the value of transaminase using the midpoint equation, where *i* now refers to the *i*^*th*^ measurement of transaminase *x* during a given acute liver injury trajectory, and further normalize *k*_*x*_ by *d*_*x*_.

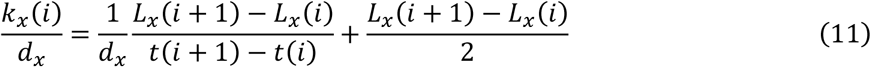

Normalization by *d*_*x*_ converts the HIX value from units of transaminase (ie, hepatocyte death) per unit time to simply units of transaminase alone; this has the effect of making the HIX directly comparable to AST and ALT measurements that clinicians are accustomed to interpreting. Thus, in Fig. 4D, when transaminase does not change from one measurement to the next, the HIX=ALT. Similarly, at steady state, Eq. 1 simplifies to *L*_*x*_*(t)=k*_*x*_*/d*_*x*_ further justifying the transformation. Substituting the plasma clearance rate *d*^*\**^_*x*_ for *d*_*x*_ in Eq. 11 produces Eq. 3. This derivation highlights that the HIX *H(t)* is asymptotically accurate in the limit of low *dt*, and progressively less accurate for larger intervals between laboratory measurements as the assumption of constancy for local *k*_*x*_ is violated. The *d*^*\**^_*x*_ was assumed to be 1.13d^-1^ for AST and 0.47d^-1^ for ALT based on earlier analyses.

## Data Availability

All data produced in the study are contained in the manuscript or on the associated github repository.

https://github.com/mssher07/LFT_trajectories_manuscript

https://hix.massgeneral.org

## List of Supplementary Materials

Materials and Methods

Figures S1 to S8

Tables S1 to S13

Data files S1 and S2

## Acknowledgments

The authors would like to thank Irun Bhan, Daniel Pratt, and Kenneth Sherman for helpful feedback about the conceptualization and application of the HIX to clinical practice.

## Author contributions

Conceptualization: MSS

Methodology: MSS

Investigation: MSS

Visualization: MSS, WG

Funding acquisition: WG

Project administration: MS, WG

Supervision: WG

Writing – original draft: MSS, WG

Writing – review & editing: MSS, WG

## Data and materials availability

All data, code, and materials used in the analysis and primers on its basic use are available at https://github.com/mssher07/LFT_trajectories_manuscript under an MIT license.

