## Supplementary Materials for "Kinetic modeling of transaminase values distinguishes active liver inflammation from resolution independent of absolute enzyme levels"

### Hepatocyte death is rarely rate-limiting among first-order kinetic trajectories

When explicitly modeling hepatocyte death, transaminase spillage into blood, and transaminase clearance, we predict that when hepatocyte death is rate-limiting, the fitted $d_{AST}$, $d_{ALT}$, and the hidden hepatocyte death rate $d_H$ will be equivalent (Fig. 2A, Fig S4. A,B). This can only occur when $d^*_{AST}$ and $d^*_{ALT}$ are sufficiently faster than $d_H$ in a particular clinical episode. We saw that indeed, among paired trajectories for $d_{AST}$ and $d_{ALT}$ (n=10,877), in the limit of low (fitted) $d_{AST}$, the ratio of $d_{AST}/d_{ALT}$ approaches 1 (Fig. 2H) consistent with the model prediction. Because these episodes are rare, and data are noisy due to variable quality of fit, we identified the threshold where hepatocyte death is rate limiting by computing confidence intervals for bins of $d_{AST}$. We define the limit where hepatocyte death is rate limiting to be the highest value of $d_{AST}$ where the 95% confidence intervals included $d_{AST}/d_{ALT} = 1$. The values and their confidence intervals are shown in Table S13, which demonstrates that the crossover point where $d_{AST}/d_{ALT} \approx 1$ occurs at values of $d_{AST} \leq 0.09$ days$^{-1}$. Using this cutoff, 29 cases were below this threshold, of 10,877 total cases. Thus, hepatocyte death is rate limiting in <0.3% of paired clinical trajectories.

### Variance minimization to estimate the plasma clearance rates of AST and ALT

In the primary analysis, $d^*_{AST}$ and $d^*_{ALT}$ are estimated by stringently filtering single-exponential fitted values of $d_{AST}$ and $d_{ALT}$ (exceptional fits, nRMSD<.015). This has the effect of asserting that only highly-first-order trajectories are included. Then, the mean of fitted $d_{AST}$ and $d_{ALT}$ were taken to be estimates of $d^*_{AST}$ and $d^*_{ALT}$. In the primary analysis, the choice of nRMSD cutoff was arbitrary and meant to assure first-order kinetics, and the cutoff was chosen based on examining the residuals of many trajectories.

A more unbiased approach to nRMSD cutoff would be to plot the nRMSD cutoffs versus the variance or standard deviation of the distributions of $d_{AST}$ and $d_{ALT}$ (Fig. S5). The variance is a result of two components, biological variability, and non-first-order kinetics. As the number of observations declines with more stringent nRMSD cutoffs, biological heterogeneity is expected to increase while non-first order kinetics continuously decreases with the more rigorous assertion of first-order kinetic fit. Therefore, the minimum of such a curve reflects a variance minimization that theoretically balances these two factors. For ALT, the minimum occurs at nRMSD<.016, and therefore yields a similar estimate as a cutoff of nRMSD<.015 (exceptional fits). For AST however, the curve continuously declines until nRMSD<.002 (n=18), and yields a slightly faster estimate ($d^*_{AST}$=1.13d$^{-1}$) compared to a cutoff of nRMSD<.015 ($d^*_{AST}$=1.05d$^{-1}$). This slightly faster estimate is visible in Fig. 2G where the moving average (black dashed line) is slightly above the exceptional-fit estimate (gray dashed line).

### Scoring liver histopathology reports for a dominant process

In the Mass General Brigham healthcare system, pathology reports are structured as a one-line summary “final diagnosis” and a “note” which provides further detail, specific stains performed, and the differential diagnosis. For simplicity and standardization, only the “final diagnosis” portion of pathology reports was considered. Pathology reports were primarily classified as indicating necrosis, hepatitis, carcinoma or normal, however if specific etiologies were explicitly stated we used these labels (e.g., graft-versus-host disease, acute cellular rejection or non-alcoholic steatohepatitis). Inflammation in the lobule (“hepatitis”) was marked as distinct from inflammation limited to the portal tracts (“portal hepatitis”). If multiple etiologies were listed, we labeled using whichever the pathologist indicated was dominant. For example, “MODERATELY ACTIVE HEPATITIS WITH PORTAL LYMPHOPLASMCYTIC INFILTRATE, PIECEMEAL NECROSIS, SPOTTY LOBULAR NECROSIS, AND BILE DUCT INJURY” would be labeled hepatitis because the stated necrosis was spotty. See Supplemental Data 1 for all final diagnoses and matched single-label dominant processes. Lastly, for the purposes of acute liver injury we considered mild or minimal steatosis, resolving liver injury, or mild to minimal cholestasis to be a variants of normal.

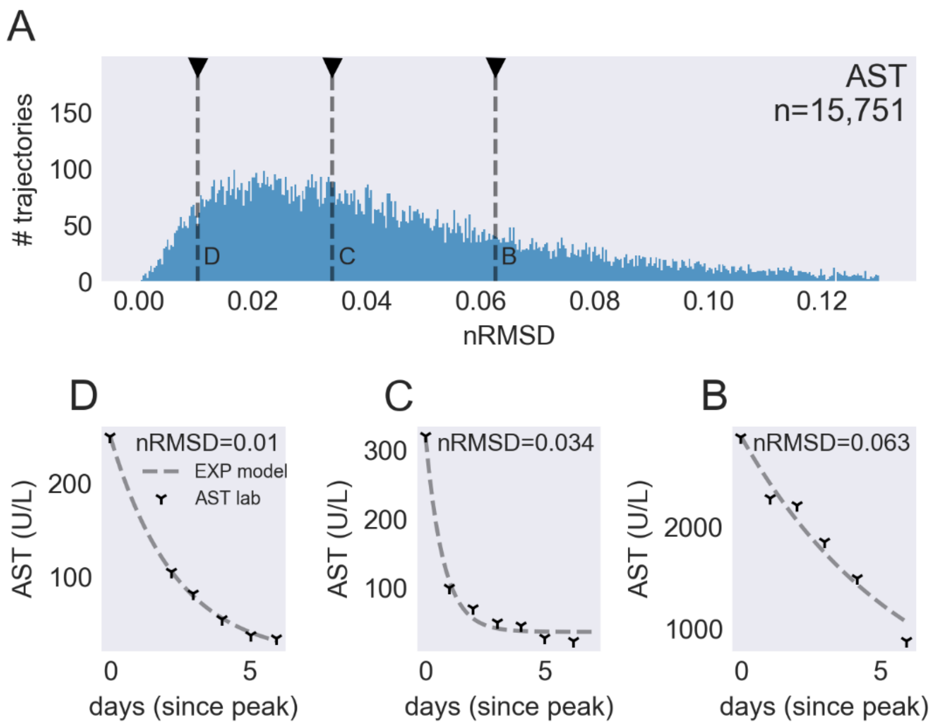


**Figure S1. Signal processing and fitting for declining AST trajectories.** (A) Distribution of goodness-of-fit in nRMSD for every declining AST trajectory in our dataset meeting criteria. Marked on the distribution are representative fits shown in sub-panels with an exceptional fit (B), an intermediate fit (C), and a poorly fit (D) trajectory.

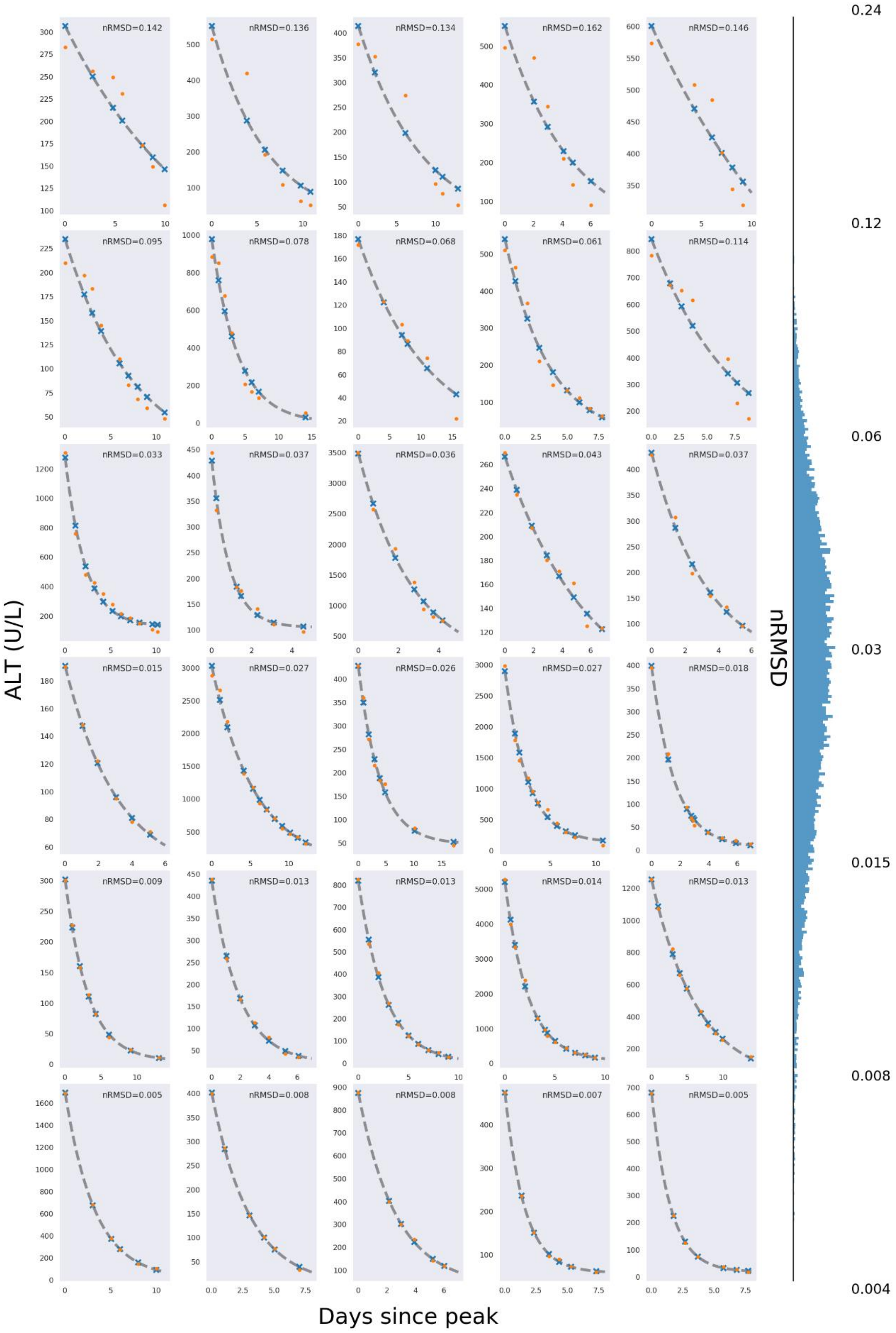

ALT (U/L)
Days since peak
nRMSD
nRMSD=0.142
nRMSD=0.136
nRMSD=0.134
nRMSD=0.162
nRMSD=0.146
nRMSD=0.095
nRMSD=0.078
nRMSD=0.068
nRMSD=0.061
nRMSD=0.114
nRMSD=0.033
nRMSD=0.037
nRMSD=0.036
nRMSD=0.043
nRMSD=0.037
nRMSD=0.015
nRMSD=0.027
nRMSD=0.026
nRMSD=0.027
nRMSD=0.018
nRMSD=0.009
nRMSD=0.013
nRMSD=0.013
nRMSD=0.014
nRMSD=0.013
nRMSD=0.005
nRMSD=0.008
nRMSD=0.008
nRMSD=0.007
nRMSD=0.005
0.24
0.12
0.06
0.03
0.015
0.008
0.004

**Figure S2. Example declining ALT trajectories by quality of goodness of fit.** On the y-axis are ALT laboratory values (orange dots) versus the fit of the simple exponential model (gray) (Eq. 2), with markers for the exact value predicted by the model at the same time point as the ALT laboratory value. The x-axis is time since ALT peak (days), and five plots are shown randomly from the dataset that have nRMSD values in the range denoted on the right. On the right, the distribution of goodness of fit values (log distributed) are plotted.

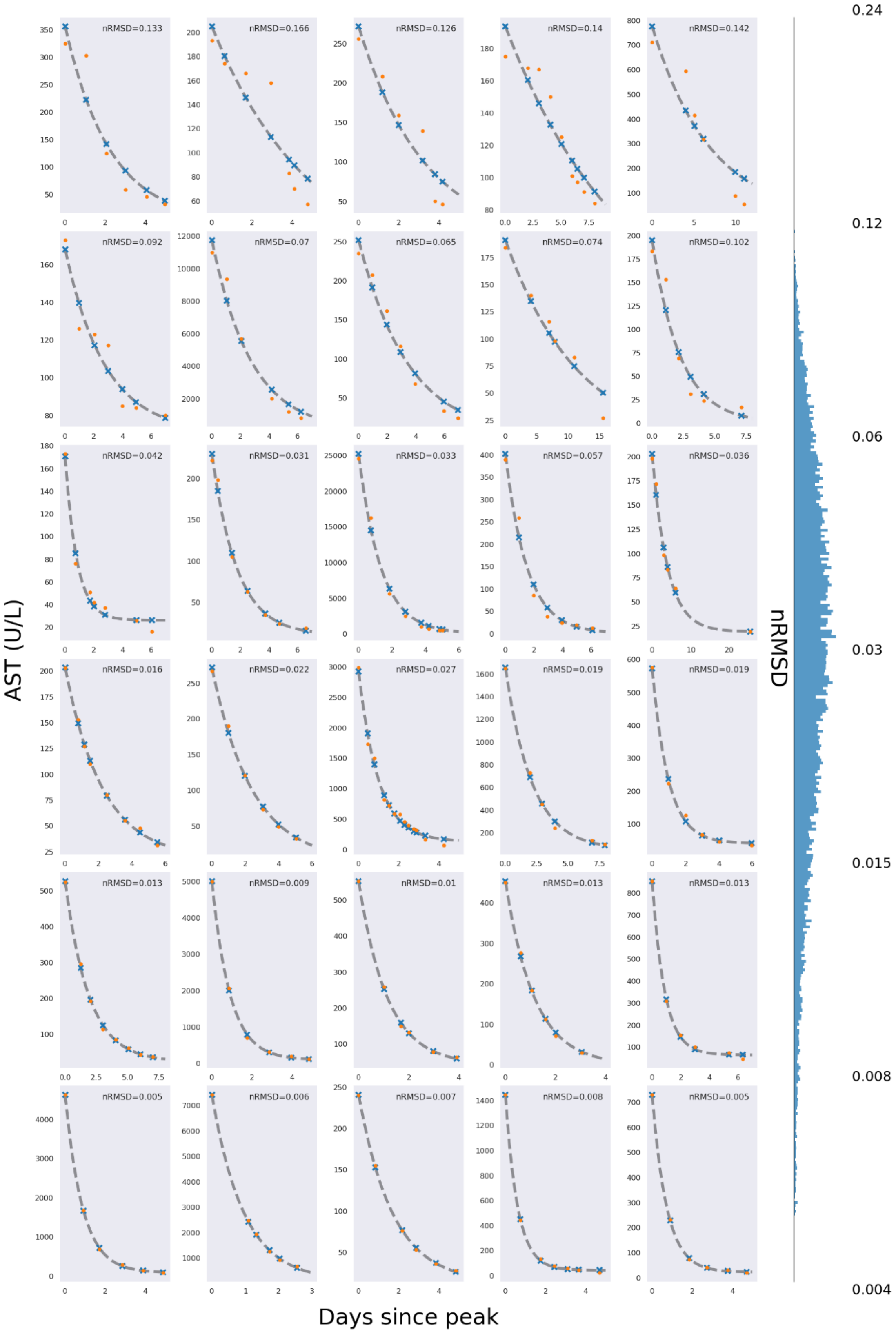
nRMSD=0.133
nRMSD=0.166
nRMSD=0.126
nRMSD=0.14
nRMSD=0.142
nRMSD=0.092
nRMSD=0.07
nRMSD=0.065
nRMSD=0.074
nRMSD=0.102
nRMSD=0.042
nRMSD=0.031
nRMSD=0.033
nRMSD=0.057
nRMSD=0.036
nRMSD=0.016
nRMSD=0.022
nRMSD=0.027
nRMSD=0.019
nRMSD=0.019
nRMSD=0.013
nRMSD=0.009
nRMSD=0.01
nRMSD=0.013
nRMSD=0.013
nRMSD=0.005
nRMSD=0.006
nRMSD=0.007
nRMSD=0.008
nRMSD=0.005
AST (U/L)
Days since peak
nRMSD
0.24
0.12
0.06
0.03
0.015
0.008
0.004

**Figure S3. Example declining AST trajectories by quality of goodness of fit.** On the y-axis are AST laboratory values (orange dots) versus the fit of the simple exponential model (gray) (Eq. 2), with markers for the exact value predicted by the model at the same time point as the AST laboratory value. The x-axis is time since AST peak (days), and five plots are shown randomly from the dataset that have nRMSD values in the range denoted on the right. On the right, the distribution of goodness of fit values (log distributed) are plotted.

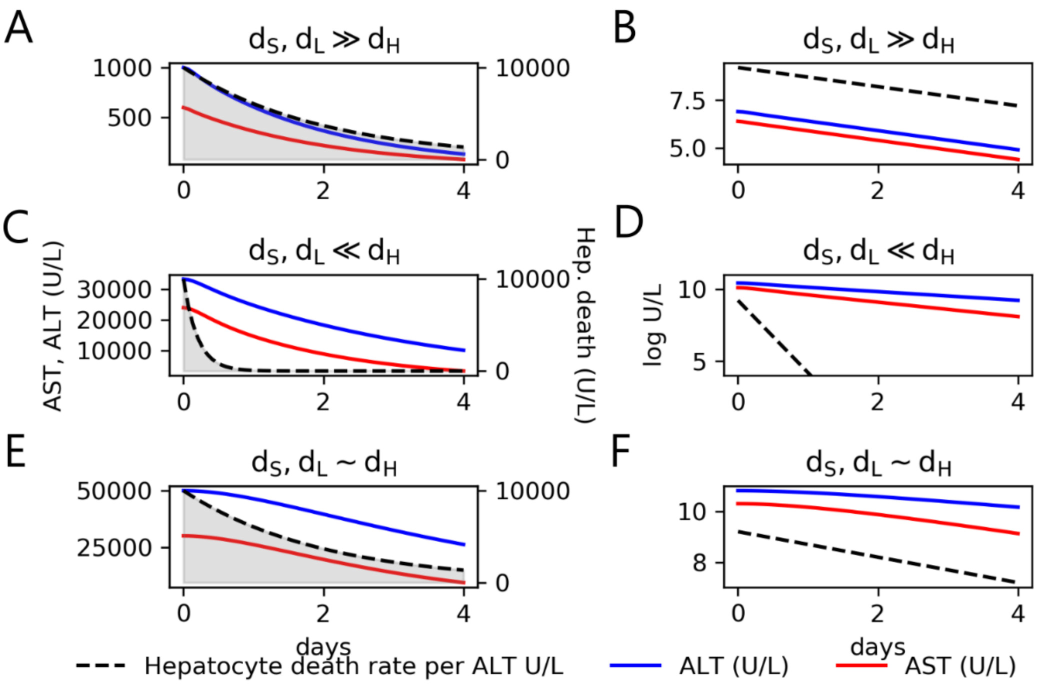


**Figure S4. Simulation of explicit models of hepatocyte death and transaminase clearance.** In (**A-F**), competing outcomes for the mechanism of a single rate-limiting step are simulated with three parameter regimes considered. On the left is plotted the trajectories of hepatocyte death resolution (dashed line, gray area) and the corresponding AST (red) and ALT (blue) curves. On the right, the log-transformed y-axis is shown for the same trajectories, where a line in the log-scaled plot indicates first-order kinetics. In (**A,B**), hepatocyte death is the rate-limiting process, which results in all three slopes in log-space (**B**) being linear with the same slope, indicating a shared rate for all three processes. In (**C,D**) hepatocyte death rapidly resolves while enzyme clearance is rate limiting, resulting in lines for all three curves in log-space (**D**) which have different slopes. Non-single-exponential behavior (**E,F**) is seen when the hepatocyte death rate, AST clearance and ALT clearance share similar rates, and leads to a curvilinear pattern in log space (**F**).

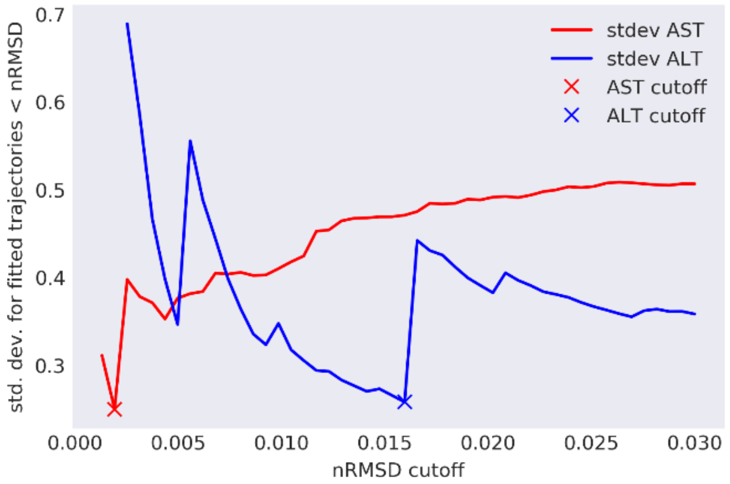


**Figure S5. Variance minimization to identify the optimal goodness-of-fit for estimating the plasma clearance rates of AST and ALT.** As goodness-of-fit (nRMSD) becomes increasingly strict (moving to the left), the number of cases included declines, but the fidelity of the model fit increases as represented by a decline in the standard deviation (variance) up to a point. The cutoff corresponding to a minima in variance corresponds to the optimal tradeoff between model fitness and case inclusion.

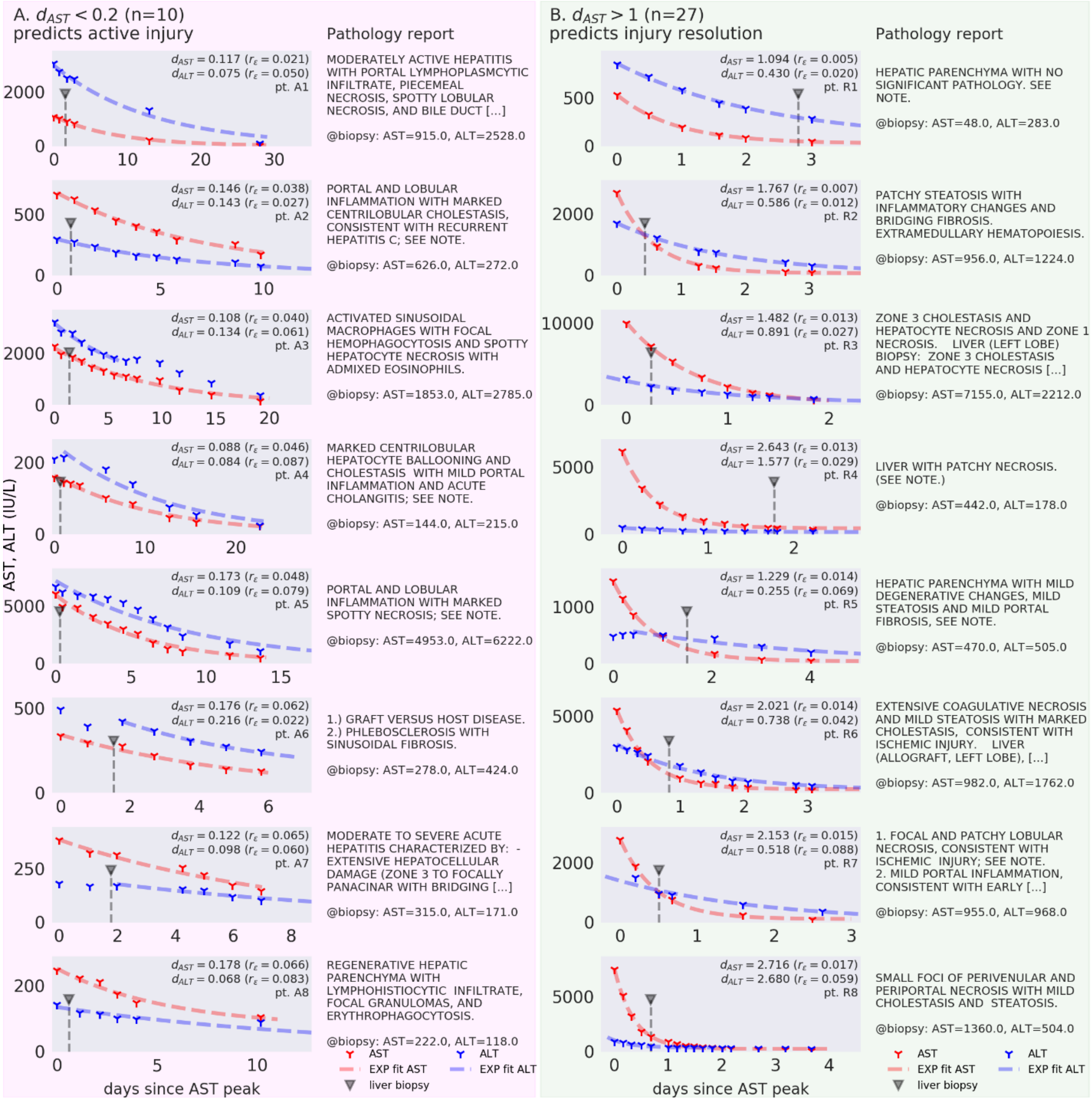


**Figure S6. Example fits of AST and ALT from the same clinical trajectory during which a liver biopsy was performed.** Each sub-figure shows a clinical episode with AST (red markers) and ALT (blue markers) decline, single-exponential model fits for each transaminase trajectory (dashed), and the time of liver biopsy (gray arrow and dashed line). In the top right corner of the plots, the value of $d_{AST}$ and $d_{ALT}$ are reported along with $r_\varepsilon$ (abbreviating nRMSD for brevity), and a patient identifier. Both (A) and (B) are sorted by best fit (smallest AST nRMSD at the top). Next to each plot is the final diagnosis text abstracted from the liver biopsy report. (A) Top 8 in descending order best-fit trajectories where $d_{AST}$ <0.2, the parameter regime where hepatocyte injury is expected to be ongoing (Fig 2A,B) and $d_{AST}$ is expected to be similar to $d_{ALT}$. (B) Top 8 in descending order best-fit trajectories where $d_{AST}$>1.0 corresponding to the parameter regime where enzyme clearance is expected to predominate (Fig 2C,D), corresponding to resolved or resolving injury.

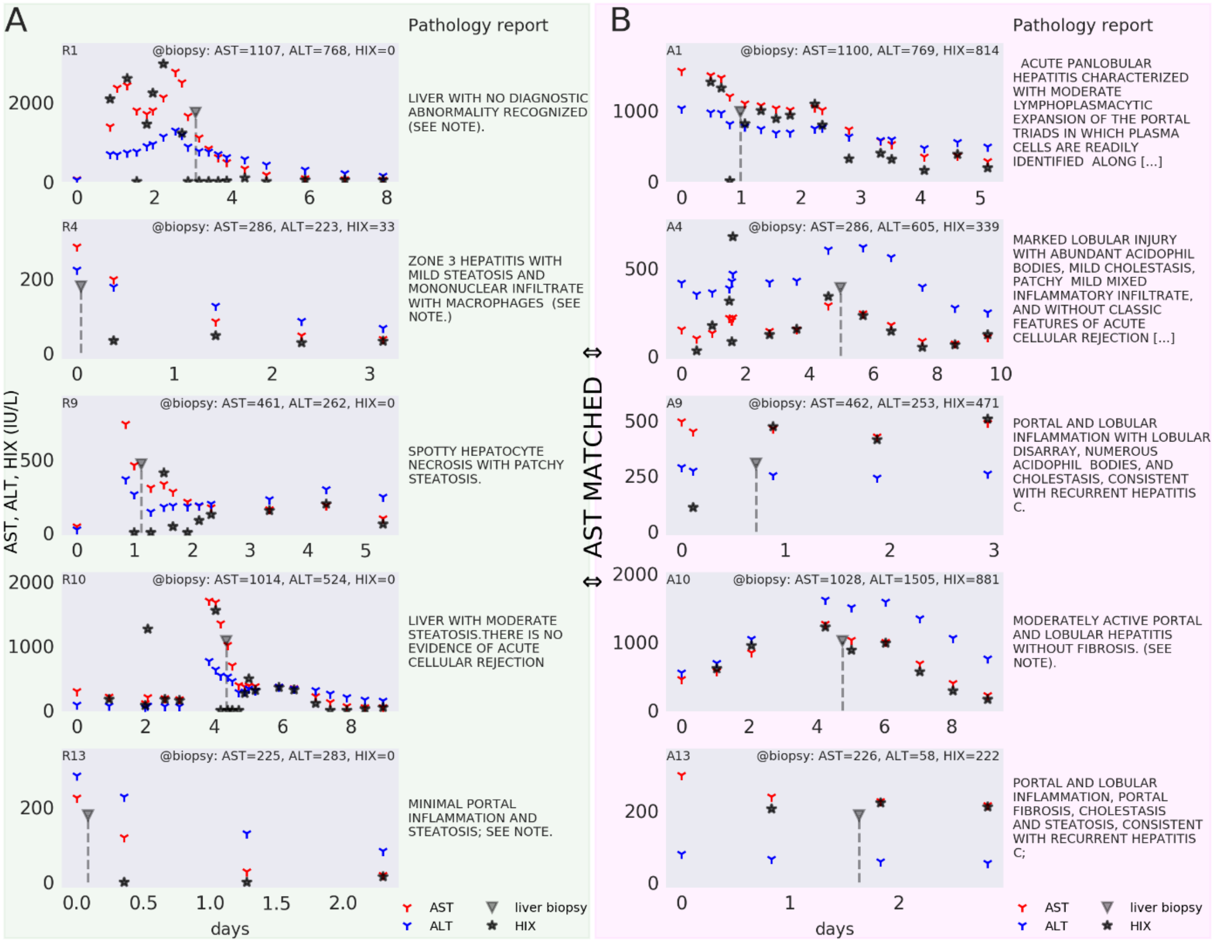


**Figure S7. HIX distinguishes active from resolved injury between trajectories when AST levels are identical or similar.** Unlike direct kinetic modeling, the HIX is calculable between any two AST values, including rising trajectories, and does not require fitting. Shown are five matched pairs from Table S9; across each row are two scenarios, one where the HIX predicts resolving injury (**A**), and a control trajectory with matched on the nearest AST to the liver biopsy where the HIX predicts ongoing injury (**B**). Each sub-figure shows a clinical episode with AST (red markers), ALT (blue markers), and HIX (black stars) calculated on consecutive AST values. The nearest AST, ALT, and HIX values to the liver biopsy are denoted in the top right corner, and the pathology final diagnosis is shown on the right.

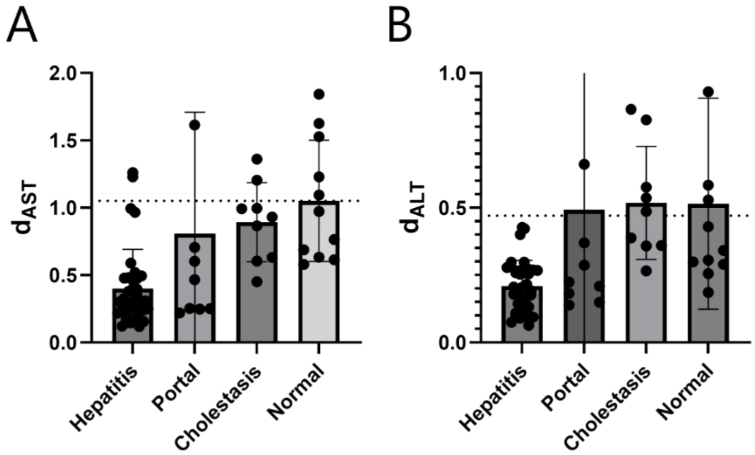


**Figure S8. Fitted $d_{AST}$ and $d_{ALT}$ correctly order four basic histopathologic processes.** Modeling predicts that lower fitted $d_{AST}$ and $d_{ALT}$ should correspond to progressively more severe disease, while higher values should correspond to progressively more normal tissue. Portal hepatitis is considered more mild ongoing injury than lobular hepatitis, and correspondingly falls above hepatitis, while cholestasis is thought to be signature of healing after recent injury, and falls between portal hepatitis and normal.

| | $d_{ALT}$ | Peak ALT (IU/L) | PT-INR | Total bilirubin | Creatinine |
|---|---|---|---|---|---|
| **$d_{ALT}$** | 1 | 0.015091 | -0.002233 | 0.026506 | 0.020188 |
| **Peak ALT (IU/L)** | 0.015091 | 1 | -0.06769 | -0.037162 | -0.049649 |
| **PT-INR** | -0.002233 | -0.06769 | 1 | 0.113295 | 0.115994 |
| **Total bilirubin** | 0.026506 | -0.037162 | 0.113295 | 1 | 0.054502 |
| **Creatinine** | 0.020188 | -0.049649 | 0.115994 | 0.054502 | 1 |

Table S1. $d_{ALT}$ correlation with clinical parameters among very good or exceptionally fit trajectories (nRMSD<.03, n= 9173)

| | $d_{AST}$ | Peak AST (IU/L) | PT-INR | Total bilirubin | Creatinine |
|---|---|---|---|---|---|
| **$d_{AST}$** | 1 | 0.067489 | -0.012459 | -0.022001 | -0.022634 |
| **Peak AST (IU/L)** | 0.067489 | 1 | -0.054099 | -0.029792 | -0.0425 |
| **PT-INR** | -0.012459 | -0.054099 | 1 | 0.105988 | 0.144859 |
| **Total bilirubin** | -0.022001 | -0.029792 | 0.105988 | 1 | 0.067401 |
| **Creatinine** | -0.022634 | -0.0425 | 0.144859 | 0.067401 | 1 |

Table S2. $d_{AST}$ correlation with clinical parameter among very good exceptionally fit trajectories (nRMSD<.03, n= 5917)

| | $d_{ALT}$ | Peak ALT (IU/L) | PT-INR | Total bilirubin | Creatinine |
|---|---|---|---|---|---|
| **$d_{ALT}$** | 1 | 0.034466 | 0.022068 | -0.007761 | 0.032667 |
| **Peak ALT (IU/L)** | 0.034466 | 1 | -0.057027 | -0.037457 | -0.036109 |
| **PT-INR** | 0.022068 | -0.057027 | 1 | 0.095126 | 0.139495 |
| **Total bilirubin** | -0.007761 | -0.037457 | 0.095126 | 1 | 0.056747 |
| **Creatinine** | 0.032667 | -0.036109 | 0.139495 | 0.056747 | 1 |

Table S3. $d_{ALT}$ correlation with clinical parameters (n= 22837)

| | $d_{AST}$ | Peak AST (IU/L) | PT-INR | Total bilirubin | Creatinine |
|---|---|---|---|---|---|
| **$d_{AST}$** | 1 | 0.068796 | 0.024926 | -0.058158 | 0.011707 |

| | | | | | |
|---|---|---|---|---|---|
| **Peak AST (IU/L)** | 0.068796 | 1 | -0.05465 | -0.045059 | -0.0368 |
| **PT-INR** | 0.024926 | -0.05465 | 1 | 0.091454 | 0.145729 |
| **Total bilirubin** | -0.058158 | -0.045059 | 0.091454 | 1 | 0.050054 |
| **Creatinine** | 0.011707 | -0.0368 | 0.145729 | 0.050054 | 1 |

Table S4. $d_{AST}$ correlation with clinical parameters (n=15751)

| | **$d_{ALT}$** | **Peak ALT (IU/L)** | **PT-INR** | **Total bilirubin** | **Creatinine** |
|---|---|---|---|---|---|
| **$d_{ALT}$** | 1 | 0.038459 | -0.020119 | 0.050655 | 0.023906 |
| **Peak ALT (IU/L)** | 0.038459 | 1 | -0.095708 | -0.032163 | -0.105317 |
| **PT-INR** | -0.020119 | -0.095708 | 1 | 0.06498 | -0.050349 |
| **Total bilirubin** | 0.050655 | -0.032163 | 0.06498 | 1 | -0.04124 |
| **Creatinine** | 0.023906 | -0.105317 | -0.050349 | -0.04124 | 1 |

Table S5. $d_{ALT}$ (nRMSD<.03) correlation with clinical parameters among patients in renal failure (Cr>2.0) (n= 2017)

| | **$d_{AST}$** | **Peak AST (IU/L)** | **PT-INR** | **Total bilirubin** | **Creatinine** |
|---|---|---|---|---|---|
| **$d_{AST}$** | 1 | 0.056947 | -0.019361 | 0.036918 | -0.000473 |
| **Peak AST (IU/L)** | 0.056947 | 1 | -0.084878 | 0.00267 | -0.077936 |
| **PT-INR** | -0.019361 | -0.084878 | 1 | 0.059637 | -0.039743 |
| **Total bilirubin** | 0.036918 | 0.00267 | 0.059637 | 1 | 0.006383 |
| **Creatinine** | -0.000473 | -0.077936 | -0.039743 | 0.006383 | 1 |

Table S6. $d_{AST}$ (nRMSD<.03) correlation with clinical parameter among patients in renal failure (Cr>2.0) (n= 1662)

| | **$d_{ALT}$** | **Peak ALT (IU/L)** | **PT-INR** | **Total bilirubin** | **Creatinine** |
|---|---|---|---|---|---|
| **$d_{ALT}$** | 1 | 0.068976 | -0.013612 | 0.059041 | -0.03578 |
| **Peak ALT (IU/L)** | 0.068976 | 1 | -0.166237 | -0.040401 | -0.073166 |
| **PT-INR** | -0.013612 | -0.166237 | 1 | 0.103776 | -0.028351 |
| **Total bilirubin** | 0.059041 | -0.040401 | 0.103776 | 1 | 0.004418 |
| **Creatinine** | -0.03578 | -0.073166 | -0.028351 | 0.004418 | 1 |

Table S7. $d_{ALT}$ (nRMSD<.03) correlation with clinical parameters among patients in liver failure (PT-INR>2.0, Total bilirubin>3.0) (n= 569)

| | $d_{AST}$ | Peak AST (IU/L) | PT-INR | Total bilirubin | Creatinine |
|---|---|---|---|---|---|
| **$d_{AST}$** | 1 | 0.111323 | 0.00563 | -0.118791 | -0.019922 |
| **Peak AST (IU/L)** | 0.111323 | 1 | -0.165082 | 0.054855 | -0.003904 |
| **PT-INR** | 0.00563 | -0.165082 | 1 | -0.013635 | 0.00948 |
| **Total bilirubin** | -0.118791 | 0.054855 | -0.013635 | 1 | -0.026745 |
| **Creatinine** | -0.019922 | -0.003904 | 0.00948 | -0.026745 | 1 |

Table S8. $d_{AST}$ (nRMSD<.03) correlation with clinical parameter among patients in liver failure (PT-INR>2.0, Total bilirubin>3.0) (n= 522)

| dominant process | n | mean_$d_{AST}$ | mean_$d_{ALT}$ | mean closest AST | 95% CI closest AST | mean closest ALT | 95% CI closest ALT |
|---|---|---|---|---|---|---|---|
| EMH | 1 | 1.767 | 0.59 | 956 | - | 1224 | - |
| NECROSIS | 33 | 1.054 | 0.65 | 1961 | 1073 | 1385 | 833 |
| NORMAL | 11 | 1.052 | 0.51 | 486 | 289 | 363 | 116 |
| CHOLESTASIS | 9 | 0.893 | 0.52 | 1412 | 1847 | 994 | 805 |
| PORTALHEP | 9 | 0.807 | 0.49 | 412 | 157 | 600 | 278 |
| CONGESTIVE | 1 | 0.804 | 0.30 | 5861 | - | 2881 | - |
| NASH | 10 | 0.609 | 0.28 | 382 | 137 | 471 | 180 |
| ACR | 6 | 0.609 | 0.32 | 633 | 321 | 821 | 309 |
| CARCINOMA | 15 | 0.524 | 0.28 | 460 | 253 | 448 | 188 |
| HEPATITIS | 35 | 0.402 | 0.21 | 1025 | 340 | 1116 | 406 |
| VOD | 3 | 0.273 | 0.21 | 604 | 525 | 490 | 166 |
| GVH | 2 | 0.251 | 0.24 | 193 | 167 | 278 | 286 |
| HLH | 2 | 0.143 | 0.10 | 1038 | 1598 | 1452 | 2614 |

Table S9. Histopathologic dominant process in comparison to model fitted $d_x$.

Table 1.

| | model | | nearest transaminases to biopsy | | | | histopathology scoring | | | | | | | |
|---|---|---|---|---|---|---|---|---|---|---|---|---|---|---|
| | HIX (IU/L) | | AST (IU/L) | | ALT (IU/L) | | normal | | lobular inflamm. | | PV/CV inflamm. | | necrosis | |
| | | | *matched* | | | | p=0.022† | | p=0.007* | | p=0.024* | | p=0.690* | |
| patient pair | resolv. | control | resolv. | control | resolv. | control | resolv. | control | resolv. | control | resolv. | control | resolv. | control |
| 1ᵠ | 0 | 814 | 1107 | 1100 | 768 | 769 | 1 | 0 | 0 | 4 | 0 | 3 | 0 | 0 |
| 2 | 0 | 236 | 350 | 351 | 420 | 891 | 0 | 0 | 1 | 1 | 2 | 3 | 2 | 0 |
| 3 | 0 | 702 | 955 | 956 | 968 | 401 | 0 | 0 | 0 | 0 | 1.5 | 0 | 1 | 1 |
| 4ᵠ | 33 | 339 | 286 | 286 | 223 | 605 | 0 | 0 | 0 | 4 | 2 | 2 | 0 | 0 |
| 5 | 0 | 1358 | 2693 | 2678 | 3589 | 1030 | 0 | 0 | 2 | 0 | 2 | 1.5 | 0 | 1 |
| 6 | 0 | 684 | 982 | 980 | 1762 | 1317 | 0 | 0 | 0 | 2 | 1 | 2 | 4 | 0 |
| 7 | 0 | 13893 | 7155 | 6946 | 2212 | 3729 | 0 | 0 | 0 | 3 | 0 | 3 | 4 | 4 |
| 8 | 0 | 1040 | 1331 | 1330 | 472 | 742 | 1 | 0 | 0 | 0 | 0 | 0 | 0 | 4 |
| 9ᵠ | 0 | 471 | 461 | 462 | 262 | 253 | 1 | 0 | 0 | 3 | 0 | 3 | 1 | 0 |
| 10ᵠ | 0 | 881 | 1014 | 1028 | 524 | 1505 | 1 | 0 | 0 | 3 | 0 | 3 | 0 | 0 |
| 11 | 0 | 4470 | 1360 | 1359 | 504 | 500 | 0 | 0 | 0 | 0 | 0 | 0 | 1 | 1 |
| 12 | 0 | 289 | 248 | 249 | 189 | 280 | 0 | 0 | 0 | 1 | 1 | 1 | 0 | 0 |
| 13ᵠ | 0 | 222 | 225 | 226 | 283 | 58 | 0 | 0 | 0 | 2 | 1 | 2 | 0 | 0 |
| 14 | 0 | 289 | 272 | 271 | 683 | 911 | 0 | 0 | 1 | 1 | 1.5 | 0 | 0 | 1 |
| 15 | 0 | 702 | 956 | 956 | 1224 | 401 | 0 | 0 | 1 | 0 | 1 | 0 | 0 | 1 |
| 16 | 0 | 391 | 400 | 400 | 481 | 323 | 0 | 0 | 0 | 0 | 0 | 2 | 4 | 0 |
| 17 | 10 | 314 | 344 | 346 | 201 | 532 | 1 | 0 | 0 | 3 | 0 | 3 | 0 | 0 |

ᵠ Plotted in Fig. S7

† Fisher's exact test

* Paired Wilcoxon signed-rank test for resolved<control

Table S10. Histopathologic dominant process in comparison to model fitted $d_x$.

| Animal | Model/measurement | AST $t_{1/2}$ | ALT $t_{1/2}$ | Reference |
|---|---|---|---|---|
| Pig | *Direct injection of AST, ALT* | | | Massarrat[1] |
| | Enzyme activity | 18h | 51h | |
| | I-131-labeling | 16h | 57h | |
| Rat | *Direct injection of AST, ALT* | | | Kamimoto[2,3] |
| | Enzyme activity, cytosolic AST | | | |
| | Fast phase | 1.9h | | |
| | Slow phase | 17.8h | | |
| | Enzyme activity, mitochondrial AST | | | |
| | Fast phase | 21min | | |
| | Slow phase | 1.5h | | |
| Rat | *Intraperitoneal injection of labeled amino acid* | | | Kim[4] |
| | Radioactive immunoprecipitated ALT | | | |
| | Fast phase | | 1.5h | |
| | Slow phase | | 84h | |
| Dog | *Direct injection of purified ALT* | | | Flescher[5] |
| | Fast phase bi-exponential model | | 17.1h | |
| | Slow phase bi-exponential model | | 61h | |
| | Mono-exponential model | | 45.2h | |
| Human | *Inference by model fit of well-behaving clinical trajectories (n=10,278)* | | | (current study) |
| | Mono-exponential model | 15.8h | 34.6h | |
| | Bi-exponential model | | | |
| | Fast phase | 26min | 24.5min, 8.8h | |
| | Slow phase | 24.4h | 56.4h | |
| | *Inference by asymptotically well-fit with variance minimization* | | | |
| | Mono-exponential model *(AST n=18, ALT n=1218)* | 14.7h | 35.1h | |
| Human | *Single-exponential model fit (n=6)* | | | Bär and Ohlendorf[6] |
| | Mono-exponential model | 17h | 47h | |
| Human | *Simple measured $t_{1/2}$ (n=5)* | | | Saheki[7] |
| | Measured half-life in clinical trajectories | | 48h | |

Table S11. Comparison of half-lives for AST and ALT in animal and human subjects.

Table S12.

| **Queried Laboratory Data (Research Patient Data Registry)** |
|---|
| ALT (SGPT) (Group:SGPT) (Loinc:1742-6) |
| ALT (SGPT) (mg/dL) (Group:ALT) (Loinc:) |
| ALT (with P5P) (Group:ALT-P5P) (Loinc:1743-4) |
| AST (SGOT) (Group:SGOT) (Loinc:1920-8) |
| Bili Conjugated (Group:CBILI) (Loinc:15152-2) |
| Bili Unconjugated (Group:UBILI) (Loinc:15153-0) |
| Bilirubin (Direct) (Group:DBILI) (Loinc:1968-7) |
| Bilirubin (Total) (Group:TBILI) (Loinc:1975-2) |
| Creatinine (Group:CRE) (Loinc:2160-0) |
| Creatinine (POC) (Group:CREAT-POC) Loinc:X1742-6) |
| PT-INR (Group:PT-INR) (Loinc:6301-6) |
| PT-INR (POC) (Group:INR-POC) (Loinc:46418-0) |

| | | $d_{AST}/d_{ALT}$ lower limit crosses 1 | | | | | | | | | | | | |
|---|---|---|---|---|---|---|---|---|---|---|---|---|---|---|
| $d_{AST}$ (days$^{-1}$) | 0.06 | **0.09** | 0.12 | 0.16 | 0.21 | 0.29 | 0.39 | 0.53 | 0.71 | 0.96 | 1.30 | 1.76 | 2.38 | 3.23 |
| $d_{AST}/d_{ALT}$ | 1.01 | 1.01 | 1.30 | 1.27 | 1.33 | 1.52 | 1.62 | 1.85 | 2.09 | 2.38 | 2.62 | 2.97 | 3.17 | 3.90 |
| $d_{AST}/d_{ALT}$ UL 95% CI | 1.17 | 1.12 | 1.48 | 1.36 | 1.40 | 1.57 | 1.66 | 1.89 | 2.13 | 2.44 | 2.69 | 3.11 | 3.51 | 5.17 |
| $d_{AST}/d_{ALT}$ LL 95% CI | 0.86 | 0.89 | 1.12 | 1.17 | 1.26 | 1.47 | 1.58 | 1.80 | 2.05 | 2.33 | 2.55 | 2.83 | 2.83 | 2.62 |

**Table S13. Values of $d_{AST}$ where $d_{AST}/d_{ALT} \approx 1$ reveals the limiting behavior where hepatocyte death is rate limiting.** The lower limit of the 95% confidence interval crosses 1 at $d_{AST}$ = 0.09.

1. Massarrat, S. Enzyme Kinetics, Half-life, and Immunological Properties of Iodine-131-labelled Transaminases in Pig Blood. *Nature* **206**, 508–509 (1965).
2. Kamimoto, Y., Horiuchi, S., Tanase, S. & Morino, Y. Plasma clearance of intravenously injected aspartate aminotransferase isozymes: Evidence for preferential uptake by sinusoidal liver cells. *Hepatology* **5**, 367–375 (1985).
3. Horiuchi, S., Kamimoto, Y. & Morino, Y. Hepatic clearance of rat liver aspartate aminotransferase isozymes: Evidence for endocytotic uptake via different binding sites on sinusoidal liver cells. *Hepatology* **5**, 376–382 (1985).
4. Kim, Y. S. The Half-Life of Alanine Aminotransferase and of Total Soluble Protein in Livers of Normal and Glucocorticoid-Treated Rats. *Mol. Pharmacol.* (1968).
5. Fleisher, G. & Wakim, K. The fate of enzymes in body fluids-an experimental study. I. Disappearance rates of glutamic-pyruvic transaminase under various conditions. *J. Lab. Clin. Med.* **61**, 76–85 (1963).
6. Bar, U. & Ohlendorf, S. Studien zur Enzymelimination. *Klin. Wochenschr.* **48**, 776–780 (1970).
7. Saheki, T. *et al.* Clearance of argininosuccinate synthetase from the circulation in acute liver disease. *Clin. Biochem.* **23**, 139–141 (1990).
